# Metabolic differences in the maternal gut microbiome precede the birth of large-for-gestational-age infants: A nested case-control study

**DOI:** 10.1101/2025.10.08.25337564

**Authors:** Anusha T. Antony, Fatemeh Mohseni, Gaël Toubon, Unnur Guðnadóttir, Lars Engstrand, Emma Fransson, Ina Schuppe-Koistinen, Nele Brusselaers, Luisa W. Hugerth

## Abstract

**Background:** Neonatal size influences newborn health and is affected by prenatal factors, such as the maternal gut microbiome, which impacts maternal metabolism and fetal growth.

**Methods:** A nested case-control study (1:2 matched) was conducted within the Swedish Maternal Microbiome (SweMaMi, 2017–2021) cohort to investigate the associations between maternal gut microbiome profiles during pregnancy and the risk of delivering large-for-gestational-age (LGA; birth weight >90th percentile) infants compared to appropriate-for-gestational-age (AGA; birth weight 10-90th percentile) infants, with a focus on dietary influence. Shotgun metagenomics enabled taxonomic and functional profiling of fecal samples collected during early (TP1: 10–20 weeks; n = 245:490) and late pregnancy (TP2: 28–32 weeks; n = 157:314). Additionally, we analyzed within-group changes in microbiome diversity from TP1 to TP2 to compare longitudinal patterns between the groups.

**Results:** At TP1, microbial species richness was higher in mothers of LGA infants (p = 0.01, 95% CI: 3.26 to 24.9), but this difference was not observed at TP2 (p = 0.30, 95% CI: –7.04 to 22.7). Beta diversity (overall microbial structure) did not differ significantly between groups at either time point. Within-sample diversity, as measured by the Shannon index, showed a significantly greater decline in the LGA group compared to the AGA group (p = 0.03, 95% CI: –0.13 to –0.007), suggesting a loss of microbial diversity specific to the LGA group over time. Evenness also declined in both groups, but the change was significantly more pronounced in the LGA group (p = 0.047, 95% CI: –0.02 to –0.0001. At TP1, the LGA group was functionally enriched in proteolytic pathways, while the AGA, group was associated with short-chain fatty acid (SCFA) production, a connection that remained at TP2. In contrast, the LGA group displayed a shift toward a pathway involved in glucose production.

**Limitations:** Reliance on self-reported dietary data, lack of physical activity and pre-pregnancy diet information, and a highly educated study population may limit generalizability.

**Conclusion:** Early pregnancy gut microbiome diversity and function, along with maternal factors, may contribute to the risk of fetal overgrowth. While microbiome diversity converges between cases and controls at a later time-point, this is not enough to mitigate risk.

## 1. Introduction

Birth weight is an important parameter for assessing a neonate’s health (1). Reference growth charts are a crucial tool for assessing the overall well-being of infants and children (2). Among several available growth charts (3), the WHO Child Growth Standards are widely regarded as robust for term infants, while the Fenton growth chart is uniquely advantageous for preterm infants. The Fenton chart provides a specific reference for monitoring growth before term age and is designed to transition seamlessly to WHO standards as the infant approaches term (4). Newborns are classified into three weight categories based on gestational age and sex according: large-for-gestational-age (LGA) if their birth weight is above the 90th percentile for gestational age; appropriate-for-gestational-age (AGA), including those between the 10th and 90th percentiles, and small-for-gestational-age (SGA) for those below the 10th percentile. A birth weight higher than 4500g, irrespective of gestational age and sex, is known as macrosomia, a condition associated with increased risk for maternal and neonatal complications (5).

The prevalence of large babies varies worldwide due to maternal health, nutrition, and genetics (4,6). Over the past 30 years, the UK, Norway, Sweden, Denmark, France, the USA, and Canada have seen rising birth weights. Globally, macrosomia and LGA occur in 3% to 15% of pregnancies (7), attributed to risk factors such as higher maternal age, increased pre-pregnancy Body Mass Index (BMI), gestational diabetes, excessive gestational weight gain, maternal depression during early pregnancy, and prenatal stress (6,8–11). Maternal complications associated with a large fetus include a higher risk of Caesarian section, postpartum hemorrhage, and perineal lacerations, which increase with the infant’s birth weight (8,12,13). LGA neonates face risks such as birth asphyxia, trauma, and hypoglycemia (12,14). In the long term, these infants may be more prone to obesity, diabetes, and cardiovascular diseases (12,13), making LGA a global health concern (13).

During pregnancy, the body undergoes significant hormonal, metabolic, and immunological changes to support fetal development (15,16). Additionally, the maternal gut microbiome also undergoes significant transformations, shifting from a more stable and comparable composition in the first trimester to more distinct changes by the third trimester (16). These transformations are typically marked by a decrease in alpha diversity and an expansion in beta diversity (16). These changes may be linked to the maternal metabolic profile. Microbial metabolites produced by the maternal gut microbiota play a crucial role in influencing the host’s metabolism during pregnancy(17). These metabolic interactions are also relevant for fetal development and the potential for pregnancy complications (17).

Recent studies suggest that maternal gut microbiome dysbiosis is linked to various pregnancy-related complications. For instance, a case-control study (15) found that women who gave birth to neonates with fetal growth restriction (FGR) had gut microbiota profiles distinct from those of healthy controls, implying a potential influence on fetal development (15). Another study (18) reported that changes in the gut microbiome can affect glucose metabolism, contributing to the onset of gestational diabetes mellitus (GDM) (18). Additionally, a study (19) profiling women in their first trimester found that those who later developed GDM exhibited altered gut microbiota, elevated pro-inflammatory cytokines, and reduced short-chain fatty acid levels months before diagnosis (19). While recent human studies have linked gut microbiota to preterm birth, more research is needed to clarify this relationship (20). A previous nested case-control study (6) conducted in pregnant women explored the maternal gut microbiome in early pregnancy in relation to macrosomia, suggesting that microbial composition and function may influence birth weight (6). Aditionally, an observational prospective cohort study (9) conducted during late pregnancy found that gut microbiota composition may play a key role in excessive fetal growth. The study hypothesized that a relatively high abundance of the Firmicutes phylum combined with a relatively low abundance of the *Prevotella* genus could be linked to LGA by influencing metabolic pathways (9). Nevertheless, the mechanistic link between specific microbial shifts and LGA outcomes remains incompletely understood, highlighting the need for more research.

Among the many factors influencing human gut microbiota, diet is one of the most significant and readily manipulated (21,22). Dietary intake during pregnancy significantly influences the maternal gut microbiome, which plays a crucial role in maternal health and fetal development (6,23). A mouse study (24) suggested that the gut microbiome may contribute to GDM, with dietary interventions potentially improving glucose tolerance by altering microbial communities (24). Additionally, an observational study (25) of 105 pregnant women revealed that diet-related factors and the gut microbiome significantly influenced postprandial glucose responses in those with diet-treated GDM (25). While the relationship between maternal diet, gut microbiome, and fetal development has been the subject of increasing research efforts, specific studies directly linking these factors to the risk of LGA are currently limited. Addressing this gap is critical given the known health implications of LGA for both mother and child.

In this nested case-control study, we compared the gut microbiome profiles of mothers who delivered large-for-gestational-age (LGA) neonates with those who delivered appropriate-for-gestational-age (AGA) neonates. We used shotgun metagenomics to investigate the taxonomic and functional profiles of the microbiome during early (TP1) and late pregnancy (TP2). Additionally, we analysed within-group changes in microbiome diversity from TP1 to TP2 (Δalpha-diversity) to compare longitudinal patterns between the groups. Finally, we assessed whether maternal dietary factors could explain the differences in their microbiome.

## 2. Materials and Methods

### 2.1 Data Collection

The study utilized samples from the Swedish Maternal Microbiome (SweMaMi) cohort, which recruited 5,423 pregnant women before 20 weeks of gestation between November 2017 and February 2021 in Sweden (26). The research was approved by the Regional Ethics Committee in Stockholm, Sweden (2017/1118-31) and conducted in accordance with international ethical guidelines. Data collection occurred at two time points (TP): gestational weeks 10–20 (TP1 or early pregnancy) and 28–32 (TP2 or late pregnancy. Participants completed online questionnaires and submitted fecal samples using at-home kits. Neonatal birth weight, gestational age at birth, and sex data were obtained from the Medical Birth Registry (26) and neonates were classified as large-for-gestational-age (LGA; birth weight > 90th percentile) or appropriate-for-gestational-age (AGA; birth weight between the 10th and 90th percentiles) using the Fenton growth chart (4). SGA infants were excluded from this study.

Participants included in this project were pregnant women who provided fecal samples and gave birth to a live infant. The primary outcome was infant gestational size at birth, grouped into LGA (case) and AGA (control) categories. The exposure was the maternal gut microbiome, assessed through fecal samples collected at two gestational time points, TP1 and TP2. Optimal 1:2 case-control matching was conducted using propensity score matching, implemented via the matchit function from the MatchIt package (27), with the matching method set to ’nearest’. Matching was based on maternal age, pre-pregnancy BMI, and gestational week at sampling.

Exclusion criteria were incomplete or missing questionnaire data, failed sample collection, subsequent pregnancies within the cohort, and sequencing errors. To assess within-group changes in gut microbial diversity (Δalpha diversity), only participants with samples available from both time points were included. Potential confounders were identified a priori and included maternal dietary factors (23) (self-reported frequency), maternal age, pre-pregnancy BMI, gestational weight gain, parity, smoking status, alcohol consumption (during pregnancy and three months before pregnancy), Perceived stress score (28), Edinburgh postnatal depression scale (29), Swedish born, antibiotics taken up to three months before pregnancy, gestational diabetes and socioeconomic status, as these may influence both the maternal microbiome and infant gestational size (6,8–11). Questionnaire data collected at TP1 and TP2 also included validated measures of demographic and lifestyle information. All variables and measurement tools are detailed in Supplementary Table S1.

### 2.2 Extraction, Sequencing and Data Annotation

Samples were collected by the participants at home, with a spoon attached to the tube’s lid and posted to Karolinska Institutet in DNA/RNA-shield buffer (Zymo Research, California, USA). Samples were kept at -80°C after arrival. For DNA extraction, samples were mechanically lysed in FastPrep-24 beads before chemical extraction with the ZymoBIOMICS 96 MagBead DNA protocol. Library preparation was performed with a brief PCR amplification step in a MGISP-960. 850-880 bp fragments were selected by electrophoresis with 4200 TapeStation. Fragments were then normalized to the same concentration and circularised. Sequencing was performed using MGI sequencers (MGI Tech Co., G400 or T7 models) on paired 150 base pair reads.

Data quality control and annotation used the StaG-mwc (v0.5.1) workflow (30) based on Snakemake (v4.8.1) (31). This workflow started with FastP (v0.23.2) (32) for trimming adapter sequences and low-quality bases. Host reads were then removed through mapping to the human reference genome GRCh38 with Kraken2 (v2.1.2) (33). Finally, the sequences were taxonomically annotated with Metaphlan4.0 (34). HUMAnN2 (HMP Unified Metabolic Analysis Network) was used for the functional analysis of metagenomic data (35). It works in conjunction with MetaPhlAn to provide functional profiling of metagenomes, gene family and pathway abundance quantification, and metabolic potential analysis of microbial communities. This allows for identifying and quantifying metabolic modules, particularly Gut Metabolic Modules (GMM) (36), in the sample data.

### 2.3 Statistical Analysis

The normality of variables was assessed by histogram. Variables are expressed as mean ± standard deviation (SD) or numbers and percentages. The comparison of continuous variables between two groups was performed by 2-tailed independent sample *t*-test. Categorical variables were compared using the χ^2^ test. All statistical analyses and plotting were done using R Statistical Software (versions 4.2.3 and 4.4.2) (37). Data was cleaned using packages from the tidyverse v2.0.0 (38) and plotted with the ggplot2 v3.5.1 (39), ggrain v0.0.4 (40). p-value < 0.05 was considered statistically significant and corrected for multiple testing with the Benjamini-Hochberg method (41).

Alpha diversity was evaluated through three key metrics: Shannon’s Diversity Index, Pielou’s Evenness, and Observed Species Richness, all calculated with the vegan package (v2.6-4/2.6.8) (42). Differences in alpha-diversity between the LGA and AGA groups were determined using a *t*-test. To assess longitudinal changes in gut microbiome diversity, within-subject differences in alpha diversity between TP1 and TP2 (Δalpha diversity) were computed. These values were compared between groups using an independent two-sample *t-*test and visualized using raincloud plots.

Beta-diversity was assessed using Jaccard, Aitchison, and Bray–Curtis distance metrics, which were computed using the ’vegdist’ function from Vegan. These distance metrics were analyzed with a permutational multivariable analysis of variance (PERMANOVA) and visualized using Principal Coordinates Analysis (PCoA) plots generated with the ggplot2 package. Aitchison distance was calculated as a robust Aitchison, which CLR-transforms values above zero. Univariable analysis was conducted for each variable, and a multivariable model was constructed using variables that were significant in the univariable analysis. Associations between dietary variables and gut microbiome alpha diversity were assessed using linear models. PERMANOVA was used as a univariate test for each variable to assess the effect of dietary factors on beta diversity.

To analyze the functional annotation data from gut microbiome samples of LGA and AGA groups, a multi-step approach was followed. Modules with a prevalence below 15% were filtered out to reduce noise and to focus on robust biological signals. The data were converted to binary format for modules with a prevalence between 15% and 85%, and either Fisher’s Exact Test or Chi-square test was applied, depending on data variance and test requirements. For high-prevalence modules (>85%), logistic regression was applied, modelling group status (Control = 0, Case = 1) as the outcome and module abundance as the predictor.

## 3. Results

### 3.1 Baseline Demographics and Descriptive Statistics

A total of 5,423 pregnant participants from the SweMaMi cohort were initially identified and classified into two groups: LGA cases (n = 305) and AGA controls (n = 2,444). After applying exclusion criteria and performing a 1:2 case-control matching, 735 participants remained at TP1, comprising 245 LGA cases and 490 AGA controls. At TP2, the matched sample included 245 LGA cases and 314 AGA controls. Figure 1 illustrates the participant selection and matching process.

**Figure 1.**
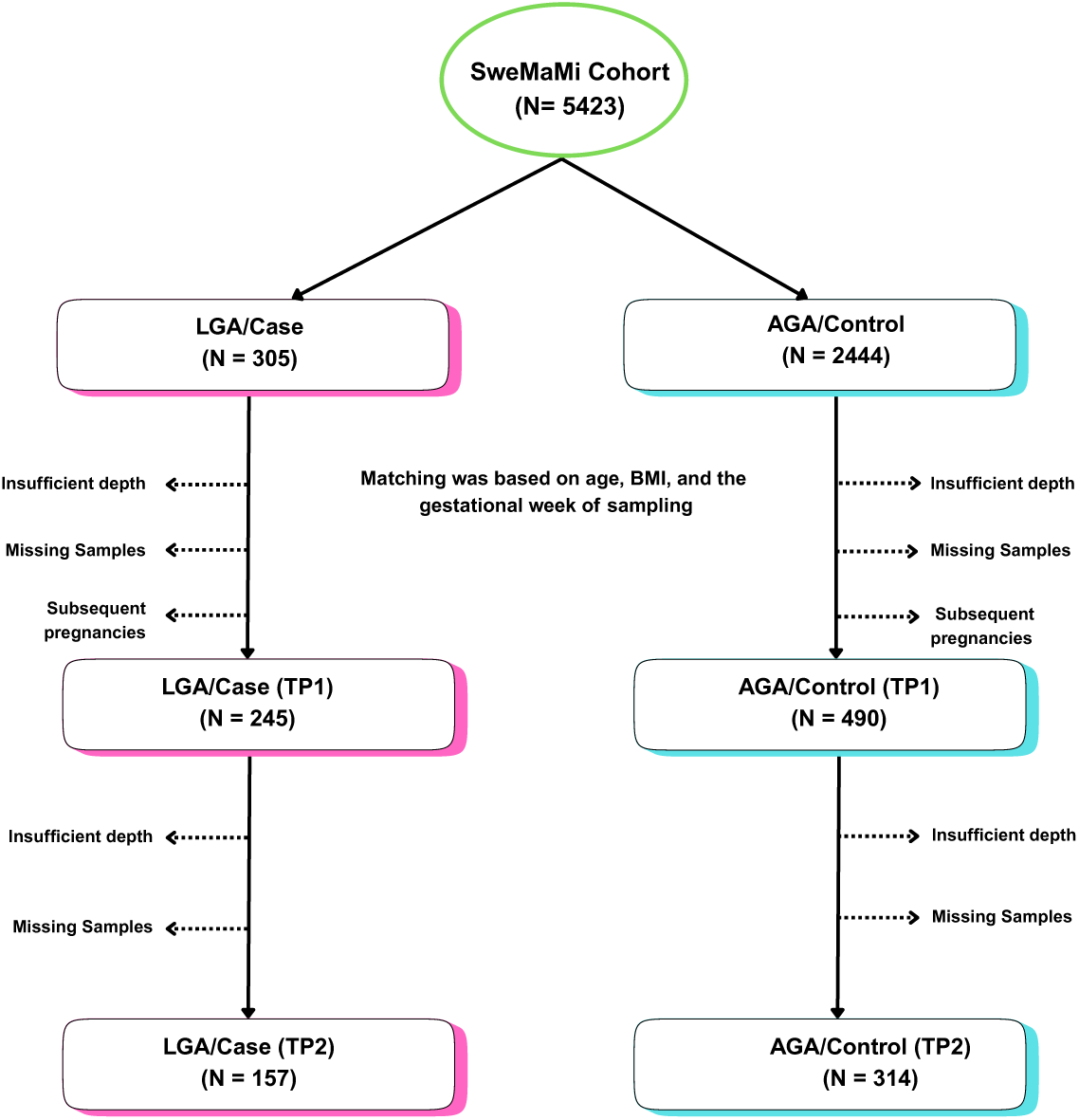
Sample selection and exclusion. N represents the number of subjects left/removed in each step. LGA: Large for Gestational Age. AGA: Appropriate for Gestational Age

The demographic and clinical characteristics of participants are presented in Table 1a for TP1 and Table 1b for TP2. The LGA group exhibited a higher prevalence of primiparity and excessive gestational weight gain during pregnancy (p < 0.05). Other variables, including maternal self-reported food frequency intakes (daily fiber, sweet drinks), smoking status, and socioeconomic status, did not differ significantly between the LGA and AGA groups (p > 0.05), as detailed in Table 1a, Table 1b, and Supplementary Table S2a, S2b.

**Table 1a.**
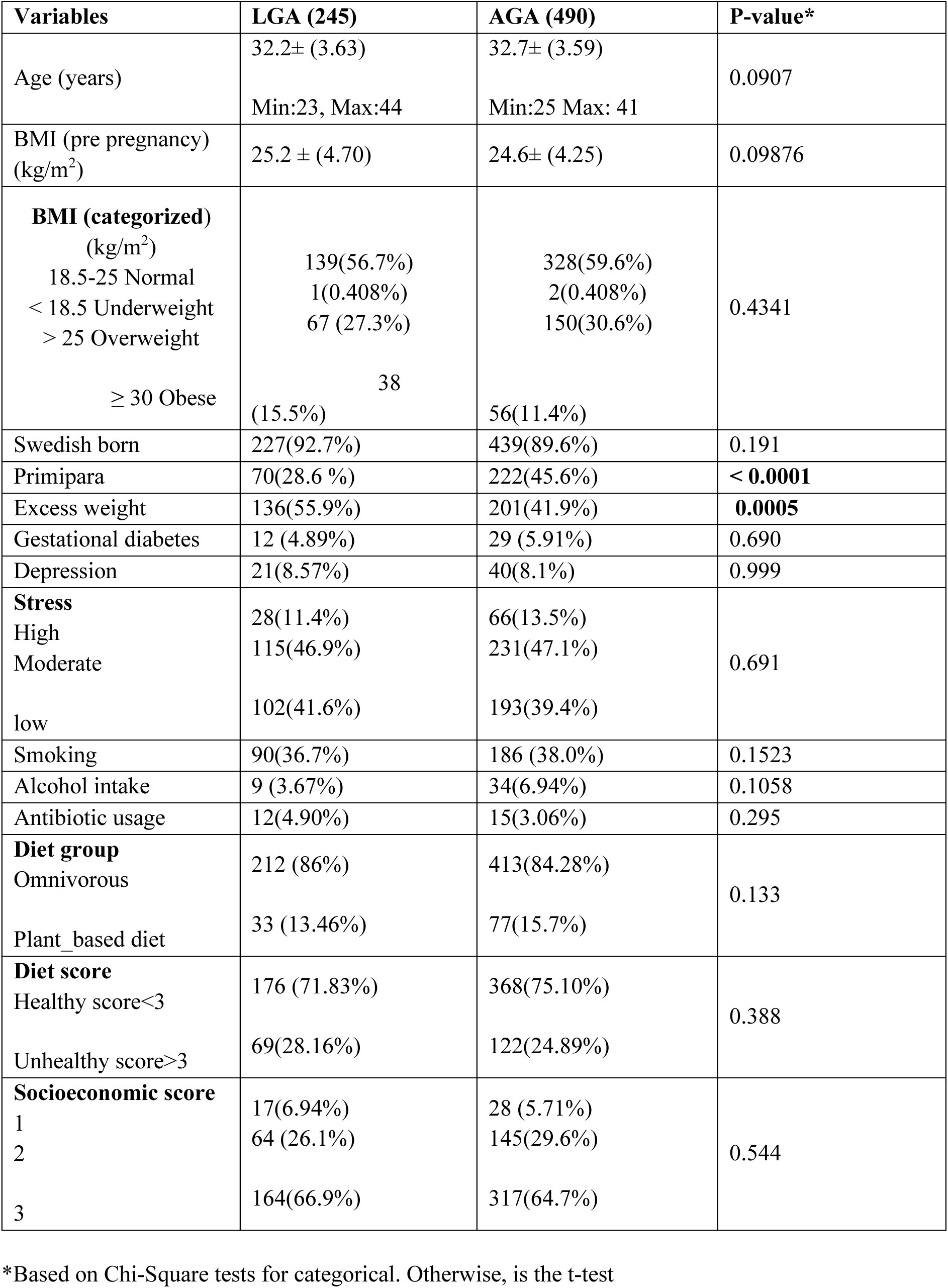
Characteristics of Study Participants for TP1.

| **Variables** | **LGA (245)** | **AGA (490)** | **P-value\*** |
| --- | --- | --- | --- |
| Age (years) | 32.2± (3.63)<br>Min:23, Max:44 | 32.7± (3.59)<br>Min:25 Max: 41 | 0.0907 |
| BMI (pre pregnancy)<br>(kg/m<sup>2</sup>) | 25.2 ± (4.70) | 24.6± (4.25) | 0.09876 |
| **BMI (categorized)**<br>(kg/m<sup>2</sup>)<br>18.5-25 Normal<br>< 18.5 Underweight<br>> 25 Overweight<br>≥ 30 Obese | 139(56.7%)<br>1(0.408%)<br>67 (27.3%)<br>38<br>(15.5%) | 328(59.6%)<br>2(0.408%)<br>150(30.6%)<br>56(11.4%) | 0.4341 |
| Swedish born | 227(92.7%) | 439(89.6%) | 0.191 |
| Primipara | 70(28.6 %) | 222(45.6%) | **< 0.0001** |
| Excess weight | 136(55.9%) | 201(41.9%) | **0.0005** |
| Gestational diabetes | 12 (4.89%) | 29 (5.91%) | 0.690 |
| Depression | 21(8.57%) | 40(8.1%) | 0.999 |
| **Stress**<br>High<br>Moderate<br>low | 28(11.4%)<br>115(46.9%)<br>102(41.6%) | 66(13.5%)<br>231(47.1%)<br>193(39.4%) | 0.691 |
| Smoking | 90(36.7%) | 186 (38.0%) | 0.1523 |
| Alcohol intake | 9 (3.67%) | 34(6.94%) | 0.1058 |
| Antibiotic usage | 12(4.90%) | 15(3.06%) | 0.295 |
| **Diet group**<br>Omnivorous<br>Plant_based diet | 212 (86%)<br>33 (13.46%) | 413(84.28%)<br>77(15.7%) | 0.133 |
| **Diet score**<br>Healthy score<3<br>Unhealthy score>3 | 176 (71.83%)<br>69(28.16%) | 368(75.10%)<br>122(24.89%) | 0.388 |
| **Socioeconomic score**<br>1<br>2<br>3 | 17(6.94%)<br>64 (26.1%)<br>164(66.9%) | 28 (5.71%)<br>145(29.6%)<br>317(64.7%) | 0.544 |

\*Based on Chi-Square tests for categorical. Otherwise, is the t-test

**Table 1b.** Characteristics of Study Participants for TP2.

| **Variables** | **LGA (157)** | **AGA (314)** | **P-value\*** |
| --- | --- | --- | --- |
| Age (years) | 32.2± (3.71)<br>Min:23, Max:44 | 32.8± (3.92)<br>Min:25 Max: 41 | 0.147 |
| BMI (pre pregnancy)<br>(kg/m<sup>2</sup>) | 25.5 ± (4.70) | 25.1 ± (4.25) | 0.244 |
| **BMI (categorized)**<br>**(kg/m<sup>2</sup>)** |  |  |  |
| 18.5-25 Normal | 83(56.7%) | 31(59.6%) | 0.402 |
| < 18.5 Underweight | - | - |  |
| > 25 Overweight | 46(27.3%) | 89(30.6%) |  |
| ≥ 30 Obese | 28(15.5%) | 32(11.4%) |  |
| Swedish born | 142(90.4%) | 230(91.3%) | 0.999 |
| Primipara | 44(28%) | 113(44.8%) | **0.0009** |
| Excess weight | 89(57.05%) | 109(44.12%) | **0.01** |
| Gestational diabetes | 10 (6.36%) | 17(6.74 %) | 0.999 |
| Depression | 6(3.82%) | 17(6.75%) | 0.219 |
| **Stress** |  |  |  |
| High | 20(12.7%) | 33(13.1%) | 0.364 |
| Moderate | 75(47.8%) | 103(40.9%) |  |
| low | 62(39.5%) | 116(46.0%) |  |
| Smoking | 62 (39.5%) | 96 (38.1%) | 0.859 |
| Alcohol intake | 6 (3.82%) | 8(3.17%) | 0.505 |
| Antibiotic usage | 17(10.8%) | 25(9.92%) | 0.963 |
| **Diet group** |  |  |  |
| Omnivorous | 137(87.26%) | 221(70.38%) | 0.623 |
| Plant_based diet | 20(12.7%) | 31(9.87%) |  |
| **Diet score** |  |  |  |
| Healthy score<3 | 112(71.33%) | 45(28.66%) | 0.899 |
| Unhealthy score>3 | 177(70.23%) | 75(29.76%) |  |
| **Socioeconomic score** |  |  |  |
| 1 | 15(9.55%) | 15(5.95%) | 0.392 |
| 2 | 46(29.3%) | 75(29.8%) |  |
| 3 | 96(61.1%) | 162(64.3%) |  |

\*Based on Chi-Square tests for categorical. Otherwise, is the t-test

### 3.2 The relationship between Gut Microbiome and LGA: Alpha and Beta diversity

Observed species richness was higher in the LGA case at TP1 (p = 0.01, Table S3, Figure 2B), but no significant differences were observed at TP2 (p = 0.30, Table S3 Figure 3B), and other alpha diversity indices did not show significant differences at either time point (p > 0.05, Figure 2, Figure 3, Table S3).

**Figure 2.**
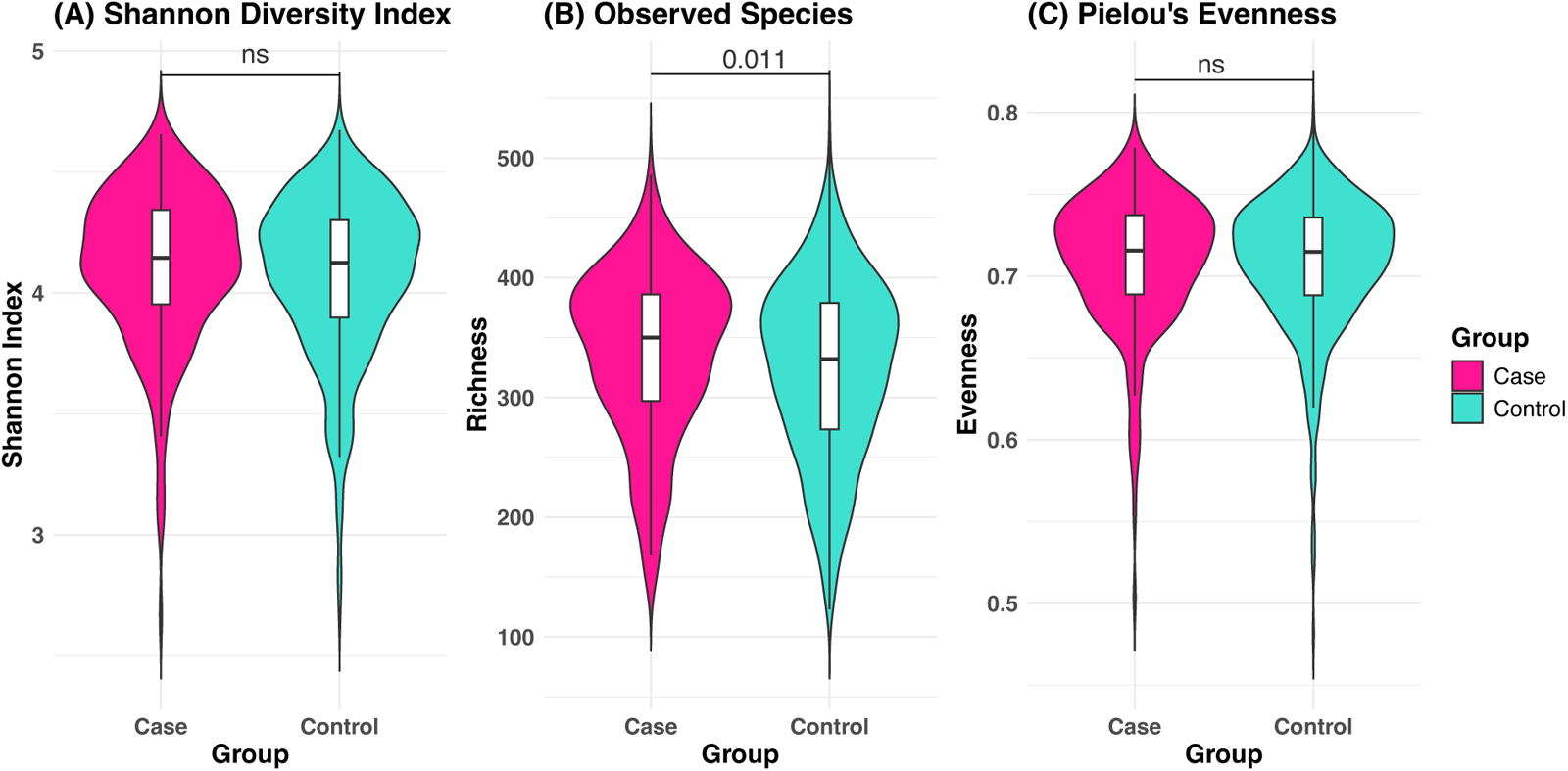
Comparison of alpha diversity between the LGA and AGA groups at TP1. Violin plot representing: A Diversity (Shannon’s diversity). B Richness (Observed species), C Evenness (Pielou’s). *P* < 0.05 indicates statistical significance, ns = not significant.

**Figure 3.**
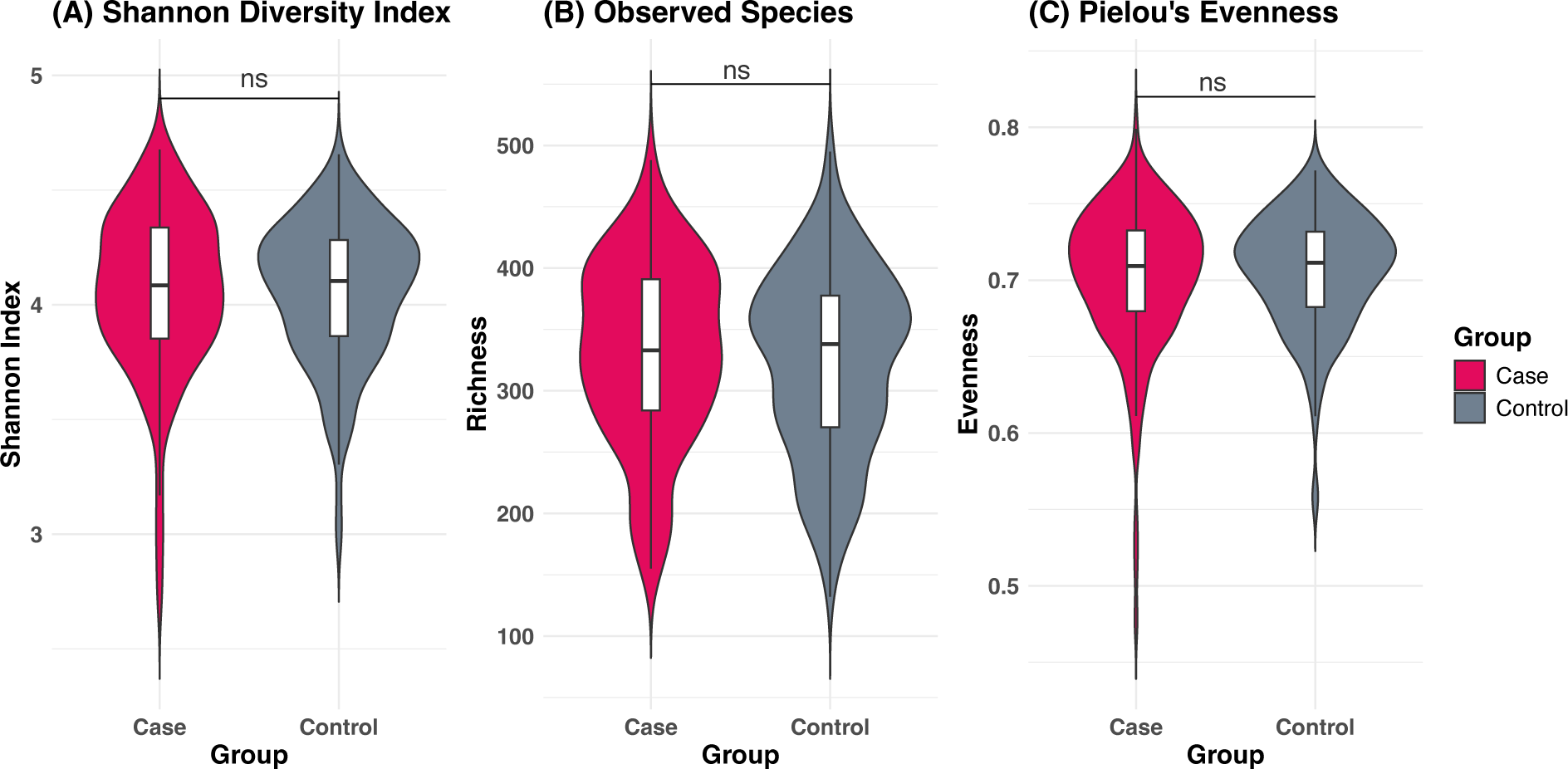
Comparison of alpha diversity between the LGA and AGA groups at TP2. Violin plot representing: A Diversity (Shannon’s diversity). B Richness (Observed species), C Evenness (Pielou’s), *P* < 0.05 indicates statistical significance, ns = not significant.

Within-subject changes in alpha diversity between TP1 and TP2 (Δalpha diversity, LGA = 157, AGA = 314), showed a substantial decline in Shannon diversity in the LGA group (p = 0.025, Table S4, Figure 4(A)), which was not observed in the AGA group. Evenness declined over time in both groups, but the change was larger in the LGA group (p = 0.047, Table S4, Figure 4 (B)). For observed species shift, the LGA group exhibited a decrease between time-points, while the AGA increased. (Table S4; Figure 4(C)). However, these changes are not statistically significant (p = 0.14).

**Figure 4.**
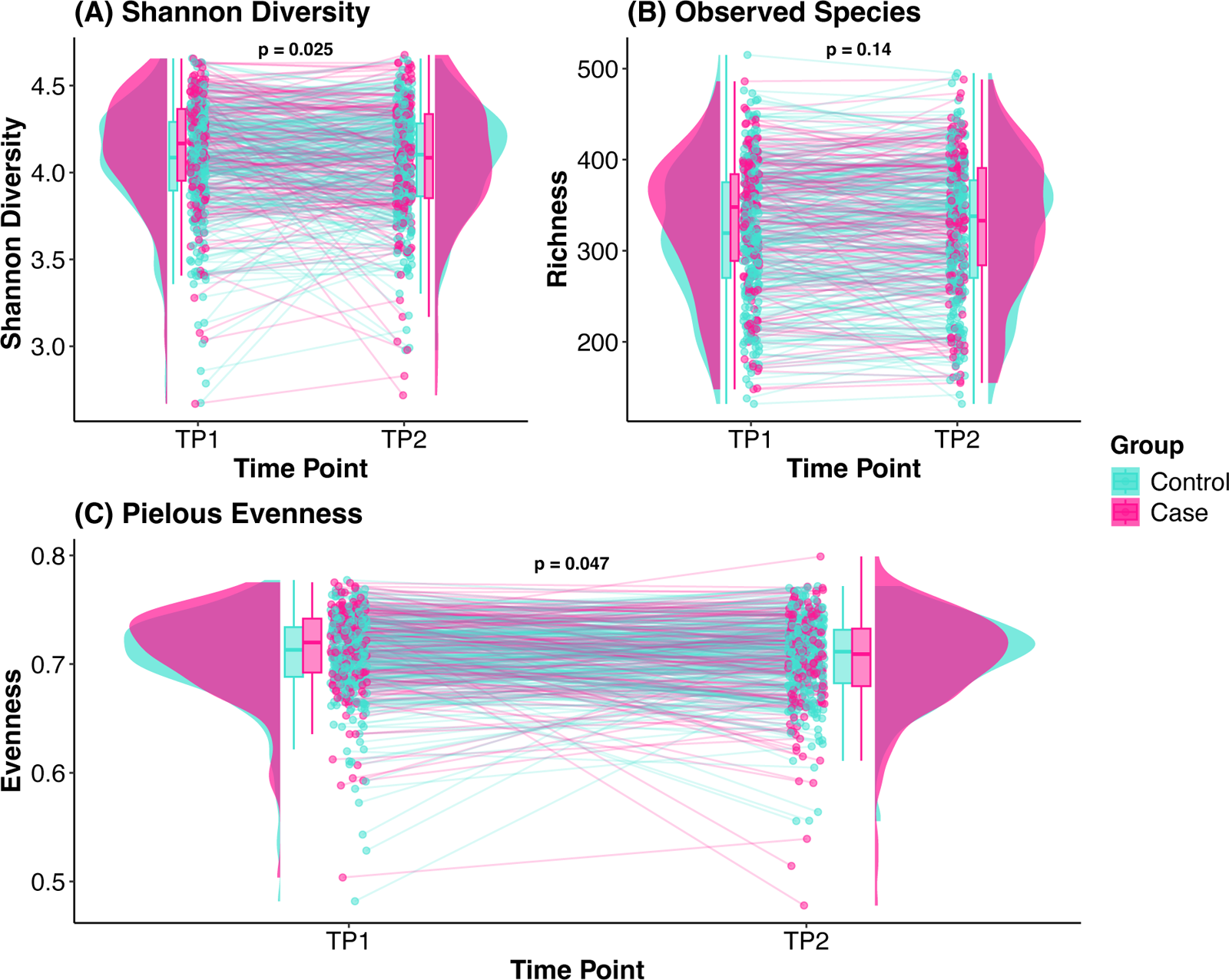
Δalpha diversity between the LGA and AGA groups: Raincloud plot representing Shannon’s diversity. B Richness (Observed species), C Evenness (Pielou’s), *P* < 0.05 indicates statistical significance.

Beta diversity was visualized through PCoA plots for TP1 (Figure 5; LGA = 245, AGA = 490) and TP2 (Figure 6; LGA = 157, AGA = 314). The Adonis test (PERMANOVA) revealed no differences in gut microbiome composition between the LGA and AGA groups at either time point. At TP1, primiparity was significantly associated with microbiome composition, as measured by Jaccard similarity, Aitchison distance, and Bray-Curtis distance (Table 2). Additionally, being born in Sweden was significantly associated with microbiome composition, as determined by Jaccard similarity Aitchison distance, and Bray-Curtis distance (Table 2). At TP2, age significantly impacted microbiome composition, as indicated by Jaccard similarity (Table 2).

**Figure 5.**
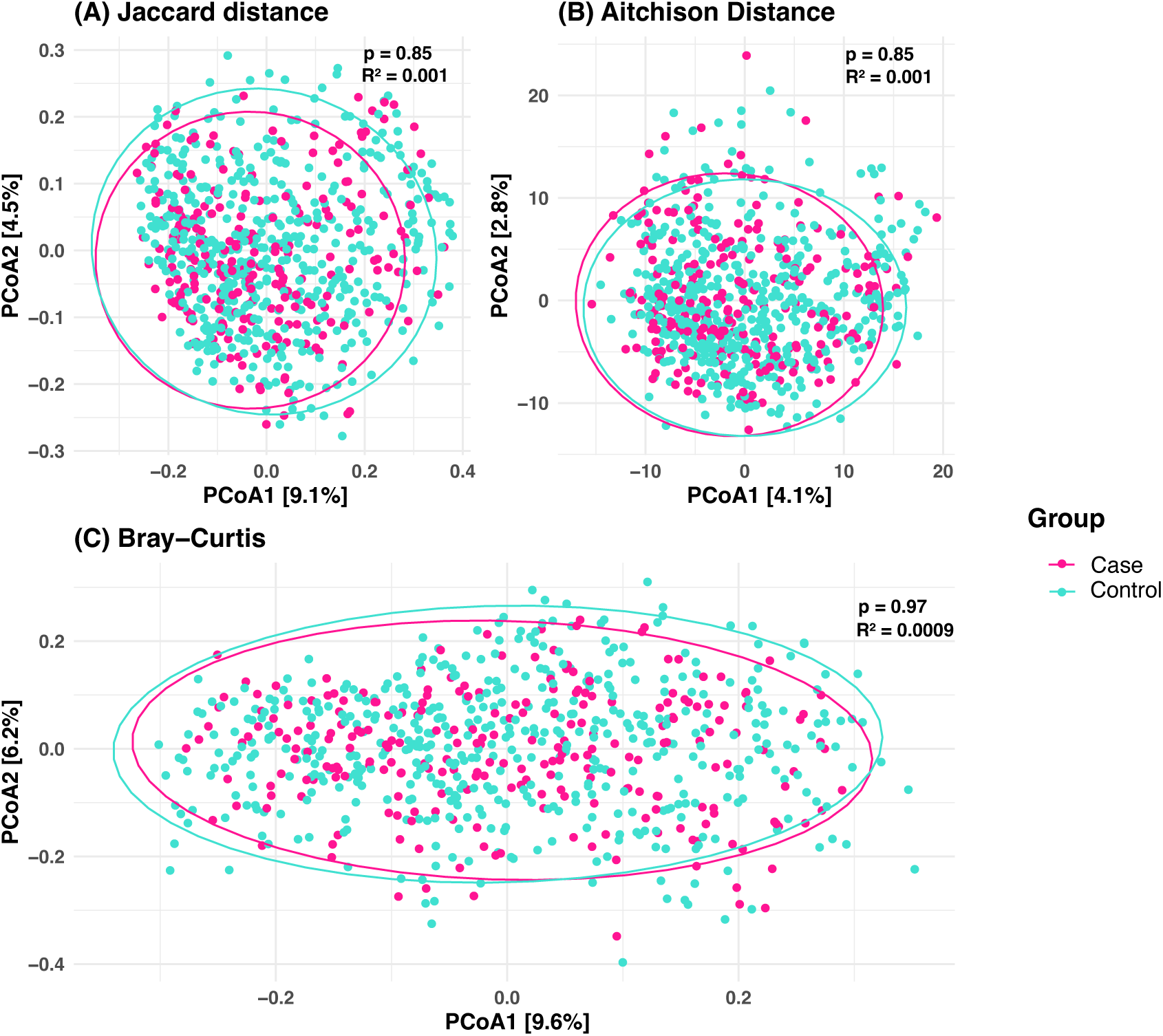
Beta Diversity at TP1: LGA vs. AGA. Plots illustrating group differences using Jaccard similarity (A), Aitchison (B), and Bray-Curtis (C) distances. P < 0.05 (PERMANOVA), R² explained by group.

**Figure 6.**
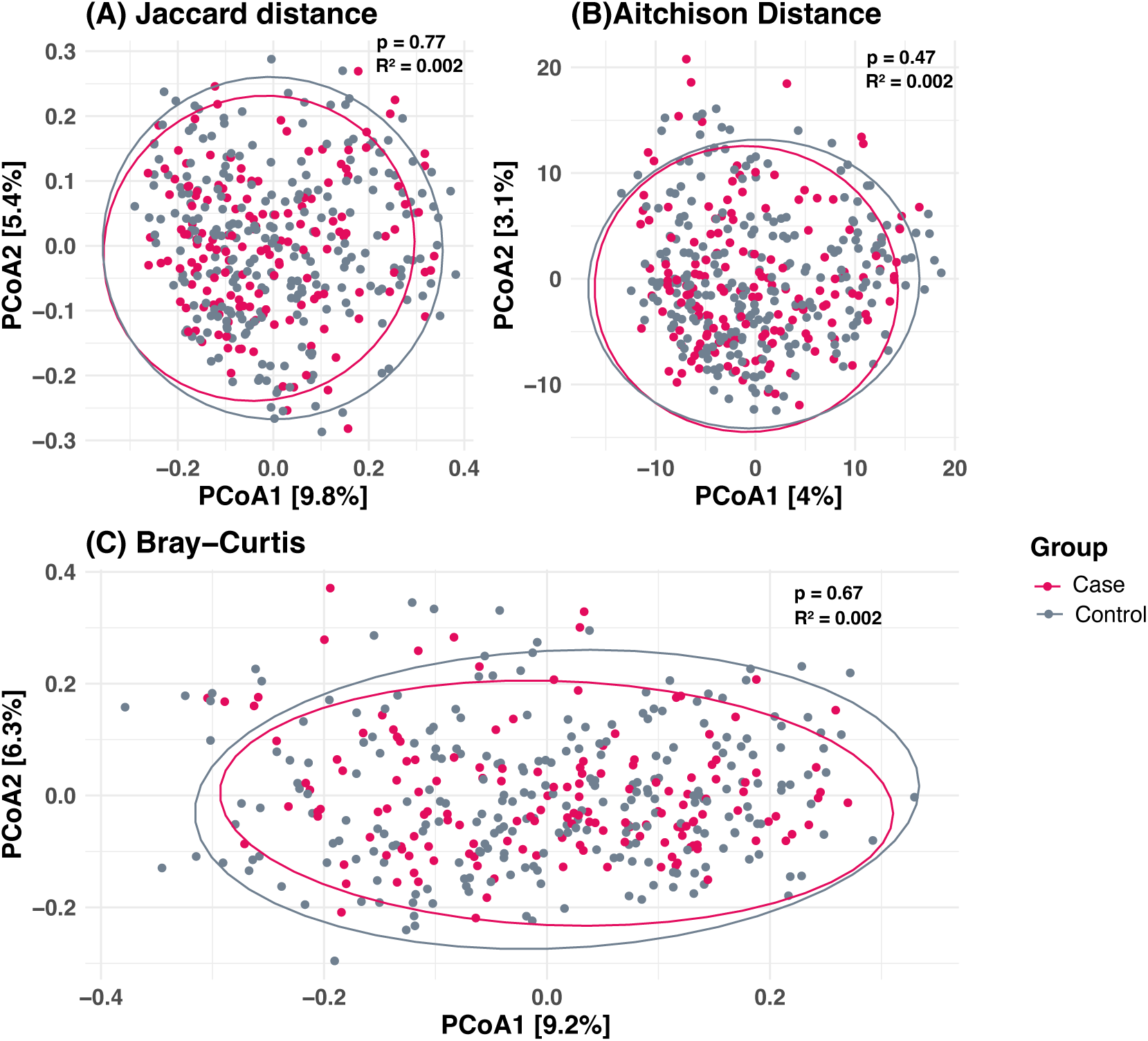
Beta Diversity at TP2: LGA vs. AGA. Plots illustrating group differences using Jaccard similarity (A), Aitchison (B), and Bray-Curtis (C) distances. p < 0.05 (PERMANOVA), R² explained by group

**Table 2.** Association between confounders and microbiome composition.

| Variables | TP1 |  |  |  |  |  | TP2 |  |  |  |  |  |
| --- | --- | --- | --- | --- | --- | --- | --- | --- | --- | --- | --- | --- |
|  | Jaccard |  | Aitchison |  | Bray-Curtis |  | Jaccard |  | Aitchison |  | Bray-Curtis |  |
|  | R<sup>2</sup> | P* | R<sup>2</sup> | P* | R<sup>2</sup> | P* | R<sup>2</sup> | P* | R<sup>2</sup> | P* | R<sup>2</sup> | P* |
| Group | 0.001 | 0.73 | 0.001 | 0.85 | 0.0009 | 0.97 | 0.002 | 0.77 | 0.002 | 0.47 | 0.002 | 0.67 |
| Maternal age | 0.001 | 0.335 | 0.001 | 0.726 | 0.001 | 0.388 | 0.003 | **0.047** | 0.002 | 0.467 | 0.002 | 0.211 |
| Pre-pregnancy BMI (categorized) | 0.003 | 0.721 | 0.004 | 0.459 | 0.0007 | 0.960 | 0.004 | 0.800 | 0.004 | 0.858 | 0.004 | 0.425 |
| Excess weight | 0.001 | 0.453 | 0.001 | 0.302 | 0.001 | 0.588 | 0.002 | 0.613 | 0.002 | 0.815 | 0.001 | 0.853 |
| parity | 0.023 | **0.002** | 0.009 | **0.015** | 0.001 | **0.040** | 0.002 | 0.722 | 0.002 | 0.668 | 0.003 | 0.129 |
| Smoking status | 0.001 | 0.355 | 0.001 | 0.625 | 0.001 | 0.212 | 0.002 | 0.479 | 0.002 | 0.966 | 0.001 | 0.830 |
| alcohol consumption (before pregnancy) | 0.001 | 0.205 | 0.001 | 0.673 | 0.001 | 0.576 | 0.002 | 0.774 | 0.002 | 0.953 | 0.002 | 0.393 |
| alcohol consumption (during pregnancy) | 0.001 | 0.316 | 0.001 | 0.427 | 0.001 | 0.680 | 0.004 | 0.928 | 0.004 | 0.974 | 0.005 | 0.354 |
| Stress score | 0.001 | 0.805 | 0.001 | 0.666 | 0.001 | 0.342 | 0.002 | 0.276 | 0.002 | 0.441 | 0.002 | 0.153 |
| Depression score | 0.001 | 0.11 | 0.001 | 0.513 | 0.001 | 0.815 | 0.002 | 0.255 | 0.003 | 0.095 | 0.002 | 0.068 |
| Swedish born | 0.002 | **0.013** | 0.009 | **0.008** | 0.001 | **0.046** | 0.002 | 0.613 | 0.002 | 0.799 | 0.002 | 0.294 |
| antibiotics taken | 0.001 | 0.066 | 0.001 | 0.676 | 0.001 | 0.309 | 0.002 | 0.128 | 0.002 | 0.115 | 0.003 | 0.125 |
| gestational diabetes | 0.001 | 0.735 | 0.001 | 0.663 | 0.0009 | 0.918 | 0.002 | 0.613 | 0.002 | 0.051 | 0.002 | 0.215 |
| socioeconomic status | 0.002 | 0.346 | 0.002 | 0.215 | 0.002 | 0.568 | 0.002 | 0.288 | 0.002 | 0.320 | 0.002 | 0.473 |

\*P-value based on Univariable PERMANOVA with adonis2. P-value <0.05 is significant.

### 3.4 Effects of Dietary Factors on Gut Microbiome Diversity and Composition

The findings indicated a positive association between daily intake of dietary fiber and vegetables and increased gut microbiome diversity at both time points (TP1 and TP2) (p < 0.05, Table 3). Specifically, fruit consumption was linked to higher species richness (p = 0.0020, Table 3) and Shannon diversity index (p = 0.049) at TP2. Although the intake of sugary and sugar-free drinks had minimal negative effects on alpha diversity (Shannon and Evenness) at both TP1 and TP2, sugary drinks at TP2 were associated with lower richness (p = 0.017, Table 3). Furthermore, unhealthy diets were strongly associated with lower microbiome diversity at both time points (Table 3). An omnivorous diet was associated with higher richness at both TP1 and TP2 (Table 3), while fish intake showed a positive association with observed species at both time points (Table 3). Other dietary variables did not demonstrate a significant association with microbiome diversity (p > 0.05). Additionally, no dietary variables showed different effects on alpha diversity between the LGA and AGA groups at both time points (p > 0.05, Supplementary Table S5). Dietary factors did not show any significant association with beta diversity at TP1 (Table 4). At TP2, the consumption of sugar-free drinks significantly impacted microbiome composition, as indicated by Jaccard similarity and Aitchison distance (Table 4).

**Table 3.**
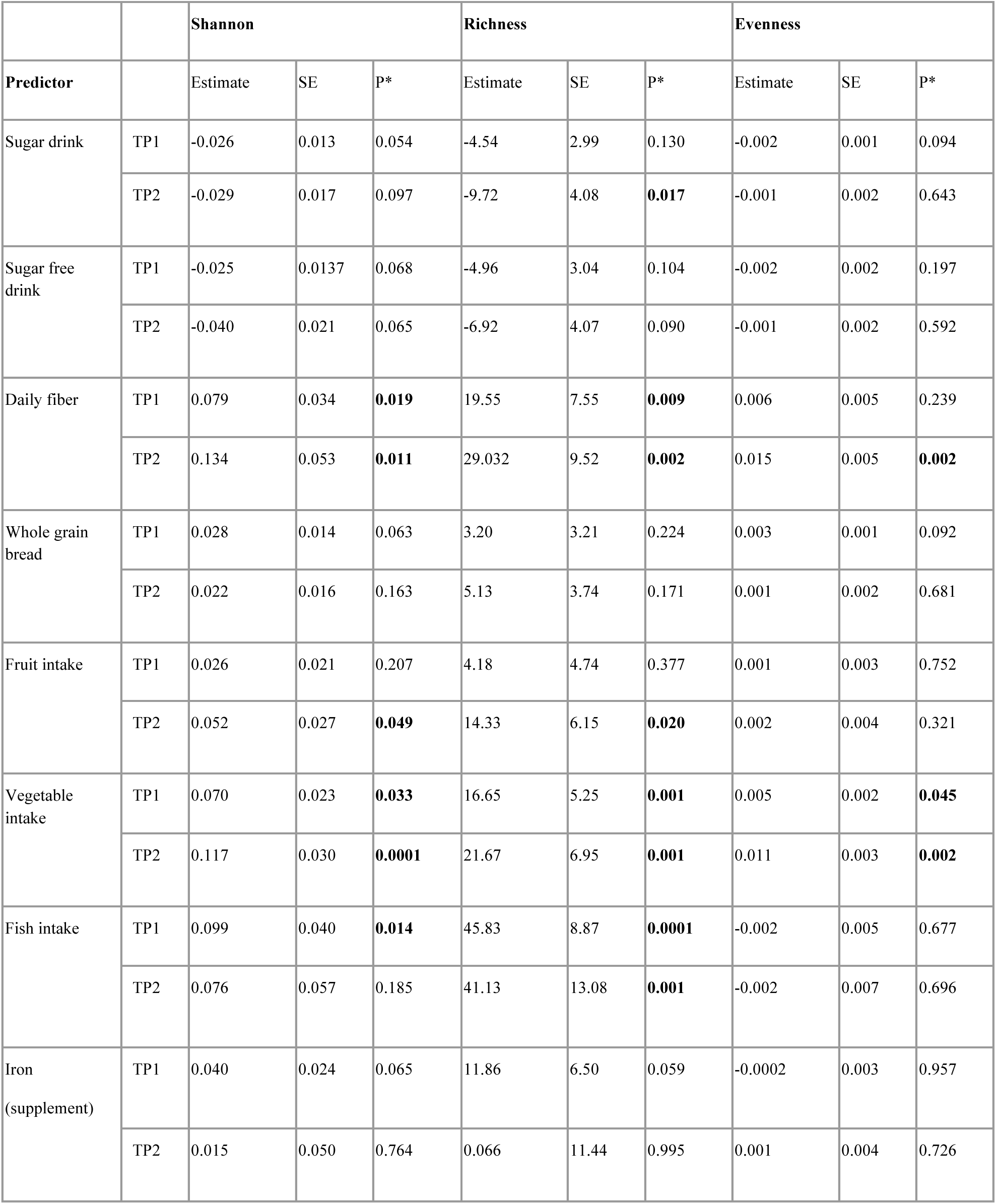
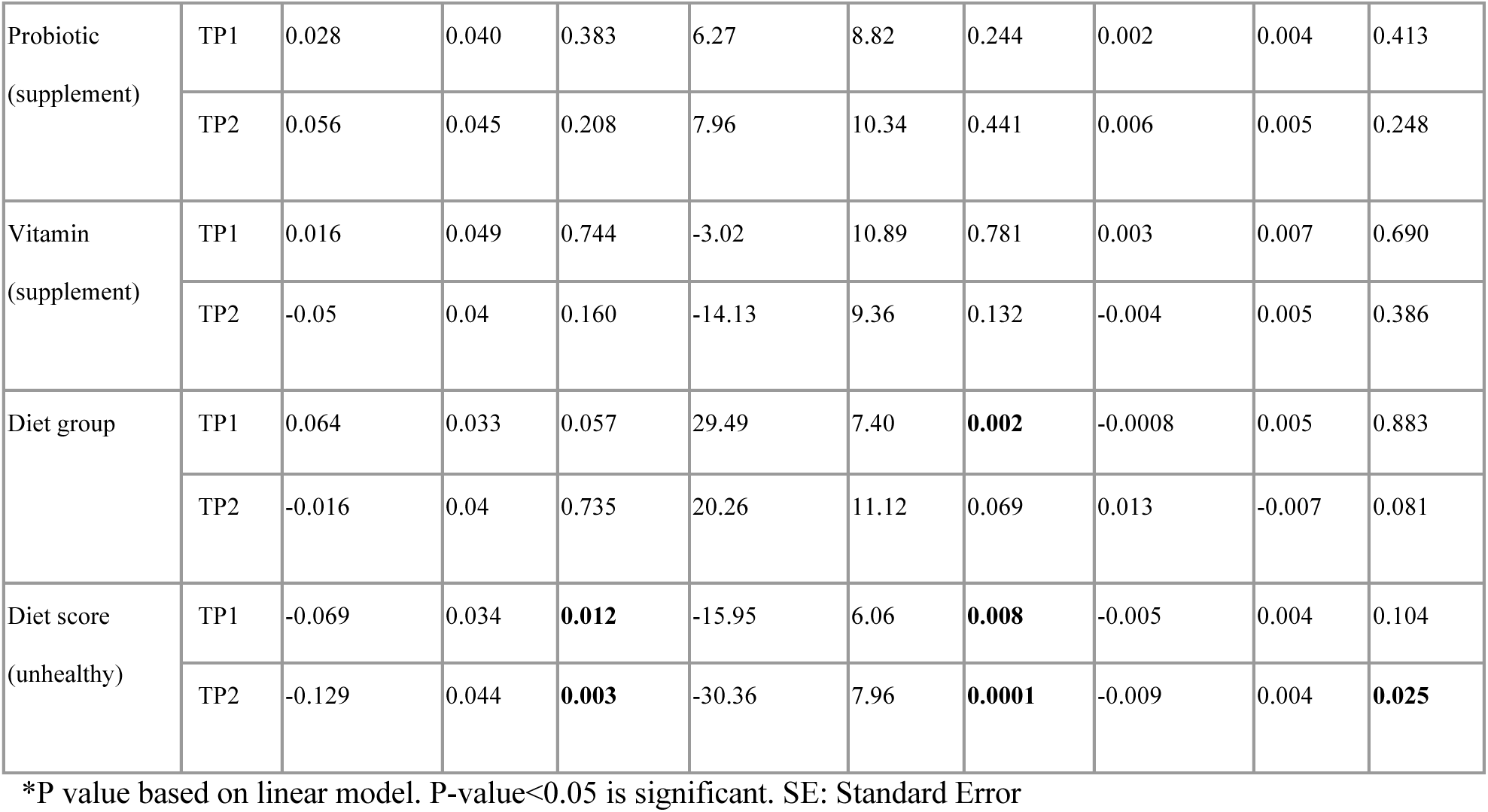
Association between dietary intake and microbiome diversity.

|  |  | Shannon |  |  | Richness |  |  | Evenness |  |  |
| --- | --- | --- | --- | --- | --- | --- | --- | --- | --- | --- |
| Predictor |  | Estimate | SE | P* | Estimate | SE | P* | Estimate | SE | P* |
| Sugar drink | TP1 | -0.026 | 0.013 | 0.054 | -4.54 | 2.99 | 0.130 | -0.002 | 0.001 | 0.094 |
|  | TP2 | -0.029 | 0.017 | 0.097 | -9.72 | 4.08 | **0.017** | -0.001 | 0.002 | 0.643 |
| Sugar free drink | TP1 | -0.025 | 0.0137 | 0.068 | -4.96 | 3.04 | 0.104 | -0.002 | 0.002 | 0.197 |
|  | TP2 | -0.040 | 0.021 | 0.065 | -6.92 | 4.07 | 0.090 | -0.001 | 0.002 | 0.592 |
| Daily fiber | TP1 | 0.079 | 0.034 | **0.019** | 19.55 | 7.55 | **0.009** | 0.006 | 0.005 | 0.239 |
|  | TP2 | 0.134 | 0.053 | **0.011** | 29.032 | 9.52 | **0.002** | 0.015 | 0.005 | **0.002** |
| Whole grain bread | TP1 | 0.028 | 0.014 | 0.063 | 3.20 | 3.21 | 0.224 | 0.003 | 0.001 | 0.092 |
|  | TP2 | 0.022 | 0.016 | 0.163 | 5.13 | 3.74 | 0.171 | 0.001 | 0.002 | 0.681 |
| Fruit intake | TP1 | 0.026 | 0.021 | 0.207 | 4.18 | 4.74 | 0.377 | 0.001 | 0.003 | 0.752 |
|  | TP2 | 0.052 | 0.027 | **0.049** | 14.33 | 6.15 | **0.020** | 0.002 | 0.004 | 0.321 |
| Vegetable intake | TP1 | 0.070 | 0.023 | **0.033** | 16.65 | 5.25 | **0.001** | 0.005 | 0.002 | **0.045** |
|  | TP2 | 0.117 | 0.030 | **0.0001** | 21.67 | 6.95 | **0.001** | 0.011 | 0.003 | **0.002** |
| Fish intake | TP1 | 0.099 | 0.040 | **0.014** | 45.83 | 8.87 | **0.0001** | -0.002 | 0.005 | 0.677 |
|  | TP2 | 0.076 | 0.057 | 0.185 | 41.13 | 13.08 | **0.001** | -0.002 | 0.007 | 0.696 |
| Iron (supplement) | TP1 | 0.040 | 0.024 | 0.065 | 11.86 | 6.50 | 0.059 | -0.0002 | 0.003 | 0.957 |
|  | TP2 | 0.015 | 0.050 | 0.764 | 0.066 | 11.44 | 0.995 | 0.001 | 0.004 | 0.726 |
| Probiotic<br>(supplement) | TP1 | 0.028 | 0.040 | 0.383 | 6.27 | 8.82 | 0.244 | 0.002 | 0.004 | 0.413 |
|  | TP2 | 0.056 | 0.045 | 0.208 | 7.96 | 10.34 | 0.441 | 0.006 | 0.005 | 0.248 |
| Vitamin<br>(supplement) | TP1 | 0.016 | 0.049 | 0.744 | -3.02 | 10.89 | 0.781 | 0.003 | 0.007 | 0.690 |
|  | TP2 | -0.05 | 0.04 | 0.160 | -14.13 | 9.36 | 0.132 | -0.004 | 0.005 | 0.386 |
| Diet group | TP1 | 0.064 | 0.033 | 0.057 | 29.49 | 7.40 | **0.002** | -0.0008 | 0.005 | 0.883 |
|  | TP2 | -0.016 | 0.04 | 0.735 | 20.26 | 11.12 | 0.069 | 0.013 | -0.007 | 0.081 |
| Diet score<br>(unhealthy) | TP1 | -0.069 | 0.034 | **0.012** | -15.95 | 6.06 | **0.008** | -0.005 | 0.004 | 0.104 |
|  | TP2 | -0.129 | 0.044 | **0.003** | -30.36 | 7.96 | **0.0001** | -0.009 | 0.004 | **0.025** |

\*P value based on linear model. P-value<0.05 is significant. SE: Standard Error

**Table 4.** Effects of dietary factors on microbiome composition.

| variables | TP1 |  |  |  |  |  | TP2 |  |  |  |  |  |
| --- | --- | --- | --- | --- | --- | --- | --- | --- | --- | --- | --- | --- |
|  | Jaccard |  | Aitchison |  | Bray-Curtis |  | Jaccard |  | Aitchison |  | Bray-Curtis |  |
|  | R<sup>2</sup> | P* | R<sup>2</sup> | P* | R<sup>2</sup> | P* | R<sup>2</sup> | P* | R<sup>2</sup> | P* | R<sup>2</sup> | P* |
| Daily fiber | 0.0012 | 0.692 | 0.0011 | 0.969 | 0.0011 | 0.700 | 0.0020 | 0.775 | 0.0024 | 0.408 | 0.0023 | 0.493 |
| Whole grain bread | 0.0038 | 0.645 | 0.0040 | 0.489 | 0.0042 | 0.324 | 0.0061 | 0.979 | 0.0073 | 0.645 | 0.0071 | 0.635 |
| Fruit intake | 0.0042 | 0.307 | 0.0041 | 0.316 | 0.0043 | 0.298 | 0.0089 | 0.078 | 0.0080 | 0.082 | 0.0090 | 0.062 |
| Vegetable intake | 0.0040 | 0.869 | 0.0040 | 0.553 | 0.0043 | 0.303 | 0.0060 | 0.976 | 0.0078 | 0.266 | 0.0065 | 0.823 |
| Sugar drink | 0.0034 | 0.941 | 0.0040 | 0.598 | 0.0038 | 0.655 | 0.0079 | 0.303 | 0.0072 | 0.731 | 0.0071 | 0.629 |
| Sugar free drink | 0.0040 | 0.484 | 0.0037 | 0.936 | 0.0041 | 0.442 | 0.0104 | **0.014** | 0.0089 | **0.003** | 0.0086 | 0.131 |
| fish | 0.0012 | 0.651 | 0.0013 | 0.553 | 0.0011 | 0.818 | 0.0022 | 0.598 | 0.0026 | 0.234 | 0.0028 | 0.239 |
| Probiotics supplement | 0.0010 | 0.953 | 0.0012 | 0.778 | 0.0011 | 0.818 | 0.0022 | 0.607 | 0.0023 | 0.632 | 0.0027 | 0.352 |
| Iron supplement | 0.0010 | 0.913 | 0.0012 | 0.903 | 0.0010 | 0.856 | 0.0038 | 0.081 | 0.0028 | 0.060 | 0.0032 | 0.095 |
| Vitamin | 0.0018 | 0.086 | 0.0016 | 0.053 | 0.0014 | 0.309 | 0.0021 | 0.721 | 0.0021 | 0.918 | 0.0019 | 0.823 |
| Diet group | 0.0012 | 0.538 | 0.0013 | 0.514 | 0.0016 | 0.149 | 0.0023 | 0.458 | 0.0022 | 0.831 | 0.0017 | 0.912 |
| Diet score | 0.0012 | 0.692 | 0.0013 | 0.627 | 0.0013 | 0.492 | 0.0022 | 0.567 | 0.0026 | 0.188 | 0.0027 | 0.257 |

\*P-value based on Univariable PERMANOVA with adonis2. P-value <0.05 is significant

### 3.5 Metabolic Module Profiling: Case-Control Comparison

At TP1, an analysis of modules with intermediate prevalence (15% - 85%) identified two input modules that were more prevalent in LGA than AGA: MF0032: Glutamate Degradation III (LGA: 69.38%, AGA: 60.40%; p = 0.0214) and MF0040: Proline Degradation (LGA: 55.5%, AGA: 44.89%; p = 0.00849) (Figure 7).

**Figure 7:**
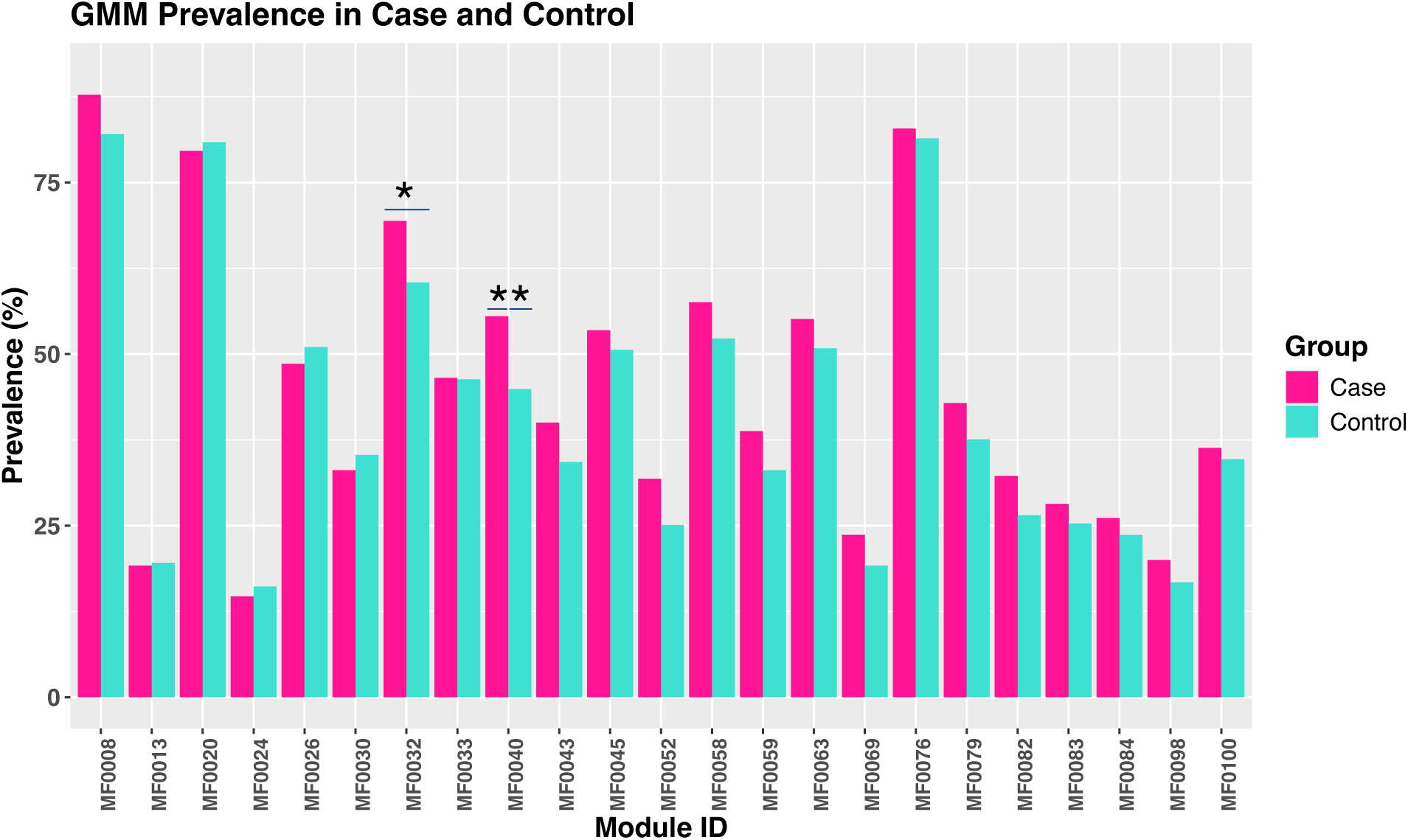
Prevalence of GMM in LGA and AGA groups at TP1. Modules with a prevalence between 15% and 85% were included. * indicate modules with statistically significant differences between the LGA and AGA groups (* = *p* < 0.05, **= p < 0.01, Chi-square test)

For high-prevalence modules (>85%), univariable logistic regression analysis revealed significant associations with LGA status. Two amino-acid degradation modules were positively associated with LGA: MF0034: alanine degradation II (OR = 1.07, 95% CI: 1.01– 1.13, p = 0.01). and MF0055: arginine degradation V (OR = 1.23, 95% CI: 1.01–1.13, p = 0.01). In contrast, several modules were negatively associated with LGA, including input modules (MF0036: isoleucine degradation and MF0041: valine degradation I), one central module (MF0073: pyruvate: ferredoxin oxidoreductase), and output modules (MF0087: acetyl-CoA to crotonyl-CoA, MF0089: butyrate production II, MF0102: sulfate reduction (dissimilatory)) (Figure 8A; Supplementary Table S6).

**Figure 8.**
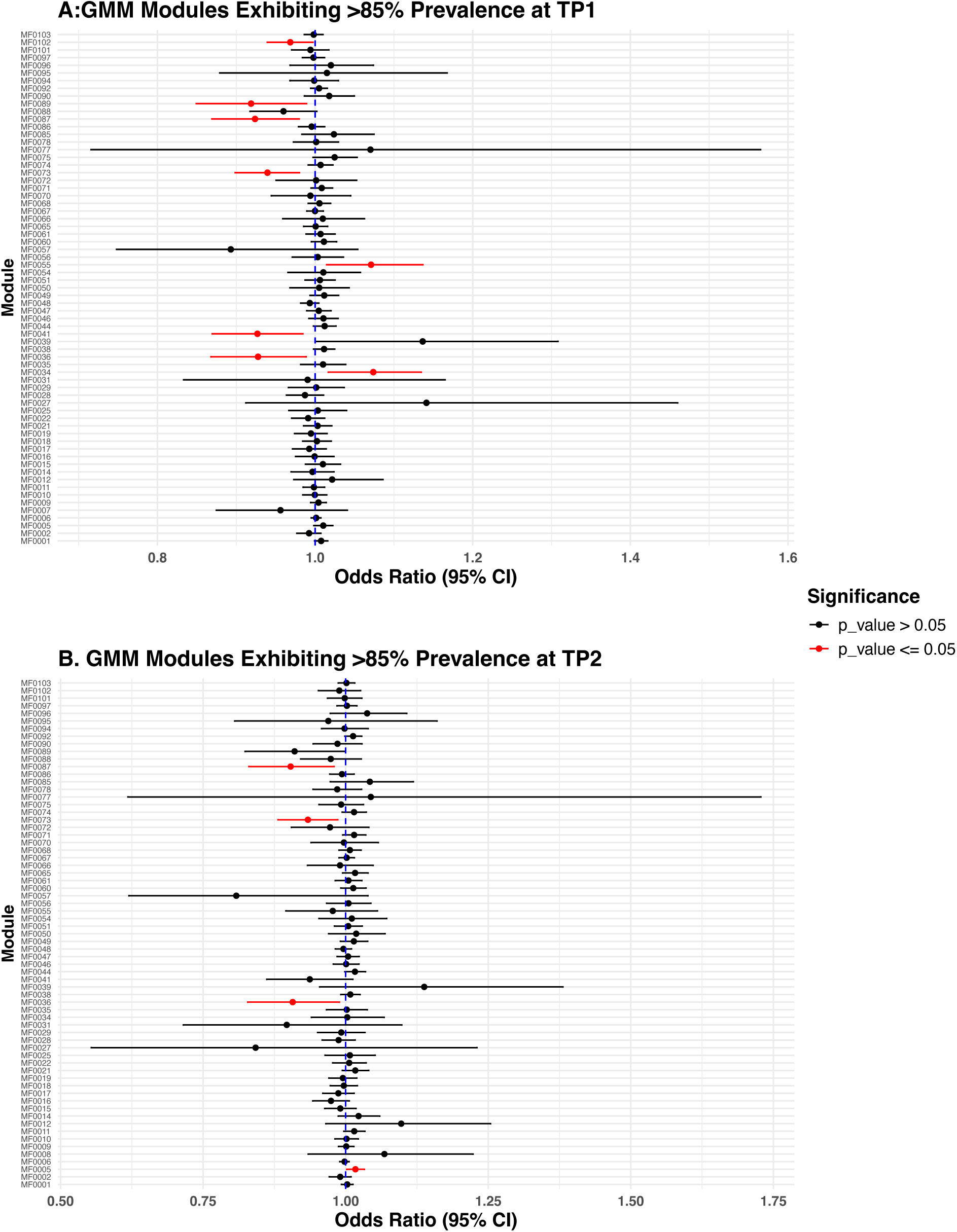
Forest Plot: Odds Ratios (OR) and 95% Confidence Intervals (CI) for High-Prevalence (>85%) Modules Associated with LGA vs. AGA at TP1(A), TP2 (B). Results from univaraible logistic regression. p-values (p < 0.05) were used to determine statistical significance.

At TP2, no modules of intermediate prevalence were significantly different (Supplementary Figure S1). However, for high-prevalence modules, there was one positive and three negative associations with LGA (Table S7; Figure 8B). The input module MF0005: starch degradation was positively associated with LGA, while the input module MF0036: isoleucine degradation was negatively associated. Additionally, the output modules MF0073: pyruvate: ferredoxin oxidoreductase and MF0087: acetyl-CoA to crotonyl-CoA exhibited negative associations with LGA group.

## 4. Discussion

This nested case-control study investigated gut microbiota composition, its longitudinal changes, and functional profiles in pregnancies leading to large-for-gestational-age (LGA) and appropriate-for-gestational-age (AGA) infants, also considering the influence of maternal diet. Our key findings reveal significant differences in gut microbiota observed species richness between LGA and AGA groups in early pregnancy, which subsequently decreased as gestation advanced. We also found that unhealthy maternal diets were strongly associated with lower microbiome diversity at both early and late pregnancy time points, with a consistent impact across both groups. Furthermore, longitudinal analysis of within-group changes indicated differences in Shannon diversity and Evenness between the groups, and functional profiling demonstrated distinct abundances of certain gut microbial metabolites (GMM) at both time points.

The LGA group at TP1 showed higher species richness (p < 0.05) than the AGA group, but no differences were observed in Shannon diversity or evenness at either time point. As pregnancy progressed, microbial species richness in the LGA group decreased, leading to no significant difference between groups at TP2. No differences in beta diversity were detected between the LGA and AGA groups. These results indicate that although alpha diversity (specifically species richness) differed at TP1, the overall microbial community structure was similar between groups. Consistent with our TP2 results, an observational cohort study of women with LGA neonates in the third trimester (9) found no statistically significant differences in overall gut microbial diversity or community composition, further supporting the idea that late pregnancy might not reveal key microbial shifts associated with LGA (6). While no studies have directly compared the maternal microbiome of mothers with LGA and AGA neonates, research on macrosomia provides useful insights. A case-control study (6) found that the overall gut microbial diversity of mothers of macrosomic babies did not significantly differ from controls.

Most existing studies have focused on late pregnancy, and data on maternal gut microbiota in early pregnancy in relation to birth weight, particularly LGA, remain scarce. Although not directly comparable, a the Women First Trial (43) assessed the maternal gut microbiome and metabolome at 12 and 34 weeks of gestation, finding that mothers of low birth weight (LBW) infants had higher microbial evenness at 12 weeks compared to those with normal birth weight (NBW) infants. This suggests early pregnancy microbial features may influence birth weight outcomes. Research on the impact of pregnancy itself on the gut microbiome is conflicting. While some studies have reported changes in either alpha or beta diversity, others have found no significant alterations . We observed notable shifts in alpha-diversity (Shannon and Evenness) from TP1 to TP2 between the AGA and LGA groups. Initially, the two groups had significant differences in gut microbial diversity, but the LGA group converged towards the diversity observed in the AGA, in line with previous SweMaMi studies . The Shannon index, reflecting both species richness and evenness, suggests these changes are tied to the abundance of certain less common taxa. Identifying these taxa could offer insights into their effects on maternal health and neonatal outcomes in LGA pregnancies.

Our analysis further examined the functional profiles of the gut microbiome at two time points, focusing on known gut-metabolic modules. All modules associated with the LGA group fall within the input module category of the GMM framework, which represents the initial steps of substrate conversion and either feeds into the central modules or drives cross-feeding interactions (36). This suggests a potential microbial functional bias toward early-stage metabolic pathways in this group. Many gut bacteria are auxotrophic, relying on cross-feeding interactions to obtain essential nutrients such as amino acids from other community members(44,45). Several LGA-associated modules reflect pathways central to microbial cross-feeding. In contrast, the metabolic modules enriched in the AGA group predominantly reflect the input, central, and output module categories.

At TP1, mothers of LGA infants had of greater proteolytic degradation potential in their guts compared to the AGA group. Proteolytic degradation is linked to microbial cross-feeding dynamics(36,45), but is also associated with low-grade inflammation (36). Some microbial metabolites can promote inflammation by influencing immune cell activation and signaling pathways (46). Inflammation during pregnancy can influence placental function, including nutrient transporter expression and activity. Changes in placental nutrient transfer efficiency may promote increased nutrient delivery to the fetus, contributing to fetal overgrowth(47). This agrees well with our finding of greater species richness in TP1, as modeling studies have found that increased frequencies of almost all amino acid auxotrophies are associated with greater microbiome diversity (44).

In contrast with the LGA group, the AGA group associated with modules involved in SCFA synthesis, a product of microbial fermentation. Notably, this includes output modules responsible for butyrate production, a key SCFA known for its multiple physiological roles. Butyrate contributes to maintaining gut epithelial integrity, modulating immune responses, and exerting anti-inflammatory effects (48). Its anti-inflammatory properties have been demonstrated in an in vitro model of preterm birth (49), highlighting its potential relevance during pregnancy. Furthermore, an in vivo mouse study (50) investigating the role of SCFAs during pregnancy found that mid-gestation SCFA supplementation increased placental weight, total placental volume, fetal vascular volume, and surface area factors that support optimal placental function and fetal development (50). Importantly, this mid-gestation window corresponds to TP1 (gestational weeks 10–20) in our cohort, aligning the findings with our study timeline. SCFAs are enriched early in pregnancy and linked to biological events like embryonic and placental development. Supplementing with probiotics or butyrate modulates placental GPR43 signaling, protecting against morphological and oxidative injury (51), suggesting a potential role for SCFAs in supporting fetal development.

Additionally, the metabolite H₂S, the product of dissimilatory sulfate reduction, is a potent signaling molecule with concentration-dependent effects: at low to moderate levels, it is anti-inflammatory and aids tissue repair in the GI tract, but excessive amounts can be toxic (52). Dietary protein intake has been positively associated with increased H₂S production in the gut (53). Although this module may be influenced by protein consumption, our questionnaire did not directly assess participants’ dietary protein intake.

All in all, the functional analysis found a pro-inflammatory profile in LGA, compared to AGA. This is specially interesting since cases and controls were matched for age and BMI, which are otherwise positively correlated with gut inflammation (54,55). Inflammation driven by specific gut microbial activities early in pregnancy may in turn alter placental nutrient transport, thereby predisposing to the delivery of an LGA infant. These findings emphasize the need to consider early gestational microbial dynamics as contributors to fetal overgrowth risk. Further studies are needed to dissect these functional-microbial interactions and their impact on fetal development.

Because the gut microbiome is strongly shaped by diet, we further assessed the association of various dietary components, including fiber, fruits, vegetables, and sugar intake, with both alpha and beta diversity of the gut microbiota. Higher fiber intake was strongly associated with microbial diversity at both time-points, in agreement with the known beneficial effects of fiber. Dietary fiber intake influences gut microbiota composition and SCFA production (56). SCFAs such as acetate, propionate, and butyrate, produced from fiber fermentation, may improve insulin sensitivity and metabolic health (9). Conversely, higher intake of sugar-sweetened beverages was associated with lower species richness at TP2. These beverages have been linked to lower microbial diversity and can negatively impact metabolic health and oxidative stress during pregnancy, potentially leading to gestational diabetes (57,58). As an alternative to sugar, artificial sweeteners are being studied for their potential to alter gut microbiota, potentially causing glucose intolerance and metabolic issues (59). Our findings suggest a link between sugar-free drinks and changes in microbial structure in late pregnancy. Additionally, fish intake was positively associated with gut microbiome diversity, supporting the Mediterranean diet’s benefits during pregnancy (23). Iron supplementation showed no significant effect on gut microbial diversity, which aligns with other studies indicating minimal impact (60). The associations observed between diet and gut microbiota in our study were not different between the LGA and AGA groups.

One of the key strengths of this study is its analysis of the gut microbiome across two critical time points during pregnancy (gestational weeks 10-20 and 28-32) on a very well characterized population. Notably, our case and control groups were matched for maternal BMI, and the prevalence of gestational diabetes was very low, especially in the case group, reducing the confounding effects of obesity and gestational diabetes, which are well-known risk factors for inflammation (61) and which lead to delivering large babies as the pregnancy outcome (62). A key limitation of this study is the use of self-reported food frequency questionnaires to assess dietary intake, which can be subject to recall bias and inaccuracies in reporting. Additionally, the assessment included only a limited number of dietary items, which may not fully represent the participants’ overall dietary patterns. Furthermore, physical activity and sleeping patterns were not collected in this study. The study population exhibited a higher level of education than the average Swedish population, which may introduce selection bias. This suggests our cohort could be more health-conscious and likely to adopt healthier eating habits. Another limitation of this study is the lack of data on dietary patterns before pregnancy. Since dietary patterns can significantly impact long-term health and microbiome diversity, the absence of information on participant’s pre-pregnancy diets limits the ability to assess any potential lasting effects on the outcomes studied. Future research would benefit from including pre-pregnancy dietary assessments to provide a more comprehensive understanding of the role of nutrition in maternal health and birth outcomes.

## 5. Conclusion

This study uniquely combines longitudinal microbiome analysis with dietary and functional assessments, providing new insights into microbiome roles in LGA risk, especially microbial diversity early in pregnancy (TP1). Longitudinally, early microbial diversity differences between the groups normalized by TP2, emphasizing the importance of early gestational monitoring. We found that functional, not overall diversity or composition, was the strongest link to fetal growth outcomes. Higher species richness in mothers of LGA infants may indicate enhanced cross-feeding and proteolytic activity, causing inflammation that could disrupt placental nutrient transport and enhance fetal growth. Conversely, SCFA synthesis pathways in the AGA group likely offer anti-inflammatory effects and support healthy development. More research is needed to identify specific bacterial species behind these differences. While diet influences overall microbial diversity, it did not differ between LGA and AGA groups, suggesting diet alone doesn’t explain these differences. Further research into additional predictors could clarify how microbiome is shaped by maternal lifestyle and influences pregnancy outcomes.

## Data Availability

All sequencing data, and limited participant-level information, can be retrieved from the European Nucleotide Archive under project ID PRJEB81814.

https://www.ebi.ac.uk/ena/browser/view/PRJEB81814

## Funding

Ferring Pharmaceuticals has funded the SweMaMi project with an unrestricted research grant. The study was also financed by SciLifeLab & Wallenberg Data Driven Life Science Program.

## Notes

### Competing Interest Statement

The authors have declared no competing interest.

### Clinical Protocols

https://pubmed.ncbi.nlm.nih.gov/36288838/

### Funding Statement

Data collection was supported by Ferring Pharmaceuticals through an unrestricted
research grant to LE. LWH and ATA are supported by the SciLifeLab & Wallenberg Data Driven Life Science Program (grant: KAW 2020.0239) to LWH

### Author Declarations

The Regional Board of Ethics, Stockholm, Sweden, gave ethical approval for this work, protocol number: 2017 1118 31

## References

1. Genowska A, Motkowski R, Strukcinskaite V, Abramowicz P, Konstantynowicz J. Inequalities in birth weight in relation to maternal factors: A population-based study of 3,813,757 live births. Int J Environ Res Public Health. 2022 Jan 26;19(3):1384.

2. Bruin C, Damhuis S, Gordijn S, Ganzevoort W. Evaluation and management of suspected fetal growth restriction. Obstet Gynecol Clin North Am. 2021 June;48(2):371–85.

3. Borghi E, de Onis M, Garza C, Van den Broeck J, Frongillo EA, Grummer-Strawn L, et al. Construction of the World Health Organization child growth standards: selection of methods for attained growth curves. Stat Med. 2006 Jan 30;25(2):247–65.

4. Fenton TR, Kim JH. A systematic review and meta-analysis to revise the Fenton growth chart for preterm infants. BMC Pediatr. 2013 Apr 20;13:59–59.

5. Damhuis SE, Ganzevoort W, Gordijn SJ. Abnormal fetal growth: Small for gestational age, fetal growth restriction, large for gestational age: Definitions and epidemiology. Obstet Gynecol Clin North Am. 2021 June;48(2):267–79.

6. Zhong Z, An R, Ma S, Zhang N, Zhang X, Chen L, et al. Association between the maternal gut microbiome and macrosomia. Biology (Basel). 2024 July 28;13(8):570.

7. Adeoye IA, Fakorede JI, Salawu MM, Adediran KI. Associations of macrosomia with sociodemographic, anthropometric, lifestyle factors and perinatal outcomes in Southwest Nigeria. BMC Pediatr. 2025 Jan 24;25(1):61.

8. McMurrugh K, Vieira MC, Sankaran S. Fetal macrosomia and large for gestational age. Obstet Gynaecol Reprod Med. 2024 Mar;34(3):66–72.

9. Lan Y, Pan S, Chen B, Zhou F, Yang F, Chao S, et al. The relationship between gut microbiota, short-chain fatty acids, and glucolipid metabolism in pregnant women with large for gestational age infants. J Appl Microbiol [Internet]. 2023 Nov 1;134(11). Available from: https://pubmed.ncbi.nlm.nih.gov/37883533/

10. Wu J, Zhou F, Xia X, Wu Y, Huang H. P-477 Associations between maternal early pregnancy depression and longitudinal fetal growth. Hum Reprod [Internet]. 2024 July 3;39(Supplement_1). Available from: 10.1093/humrep/deae108.820

11. Mélançon J, Bernard N, Forest J-C, Tessier R, Tarabulsy GM, Bouvier D, et al. Impact of maternal prenatal psychological stress on birth weight. Health Psychol. 2020 Dec;39(12):1100–8.

12. Said AS, Manji KP. Risk factors and outcomes of fetal macrosomia in a tertiary centre in Tanzania: a case-control study. BMC Pregnancy Childbirth. 2016 Aug 24;16:243.

13. Zheng J, Xiao X-H, Zhang Q, Mao L-L, Yu M, Xu J-P, et al. Correlation of placental microbiota with fetal macrosomia and clinical characteristics in mothers and newborns. Oncotarget. 2017 Oct 10;8(47):82314–25.

14. Ju H, Chadha Y, Donovan T, O’Rourke P. Fetal macrosomia and pregnancy outcomes. Obstet Anesth Dig. 2010 Dec;30(4):231–2.

15. Xiao Y, Li M, Zheng S, Pan X, Peng Y, Ning P, et al. Alterations in maternal-fetal gut and amniotic fluid microbiota associated with fetal growth restriction. BMC Pregnancy Childbirth. 2024 Nov 8;24(1):728.

16. Koren O, Goodrich JK, Cullender TC, Spor A, Laitinen K, Bäckhed HK, et al. Host remodeling of the gut microbiome and metabolic changes during pregnancy. Cell. 2012 Aug 3;150(3):470–80.

17. Lu X, Shi Z, Jiang L, Zhang S. Maternal gut microbiota in the health of mothers and offspring: from the perspective of immunology. Front Immunol. 2024 Mar 13;15:1362784.

18. Wang J, Zheng J, Shi W, Du N, Xu X, Zhang Y, et al. Dysbiosis of maternal and neonatal microbiota associated with gestational diabetes mellitus. Gut. 2018 Sept;67(9):1614–25.

19. Pinto Y, Frishman S, Turjeman S, Eshel A, Nuriel-Ohayon M, Shrossel O, et al. Gestational diabetes is driven by microbiota-induced inflammation months before diagnosis. Gut. 2023 May;72(5):918–28.

20. Di Simone N, Santamaria Ortiz A, Specchia M, Tersigni C, Villa P, Gasbarrini A, et al. Recent insights on the maternal Microbiota: Impact on pregnancy outcomes. Front Immunol. 2020 Oct 23;11:528202.

21. Mandal S, Godfrey KM, McDonald D, Treuren WV, Bjørnholt JV, Midtvedt T, et al. Fat and vitamin intakes during pregnancy have stronger relations with a pro-inflammatory maternal microbiota than does carbohydrate intake. Microbiome. 2016 Oct 19;4(1):55.

22. Amato KR, Pradhan P, Mallott EK, Shirola W, Lu A. Host-gut microbiota interactions during pregnancy. Evol Med Public Health. 2024 Jan 6;12(1):7–23.

23. Maher SE, O’Brien EC, Moore RL, Byrne DF, Geraghty AA, Saldova R, et al. The association between the maternal diet and the maternal and infant gut microbiome: a systematic review. Br J Nutr. 2023 May 14;129(9):1491–9.

24. Frishman S, Nuriel-Ohayon M, Turjeman S, Pinto Y, Yariv O, Tenenbaum-Gavish K, et al. Positive effects of diet-induced microbiome modification on GDM in mice following human faecal transfer. Gut. 2024 Sept 9;73(10):e17.

25. Popova PV, Isakov AO, Rusanova AN, Sitkin SI, Anopova AD, Vasukova EA, et al. Personalized prediction of glycemic responses to food in women with diet-treated gestational diabetes: the role of the gut microbiota. NPJ Biofilms Microbiomes. 2025 Feb 7;11(1):25.

26. Fransson E, Gudnadottir U, Hugerth LW, Itzel EW, Hamsten M, Boulund F, et al. Cohort profile: the Swedish Maternal Microbiome project (SweMaMi) - assessing the dynamic associations between the microbiome and maternal and neonatal adverse events. BMJ Open. 2022 Oct 26;12(10):e065825.

27. Ho DE, Imai K, King G, Stuart EA. MatchIt: Nonparametric Preprocessing for Parametric Causal Inference. J Stat Softw [Internet]. 2011;42(8). Available from: 10.18637/jss.v042.i08

28. Cohen S, Kamarck T, Mermelstein R. A global measure of perceived stress. J Health Soc Behav. 1983 Dec;24(4):385–96.

29. Cox JL, Holden JM, Sagovsky R. Detection of postnatal depression. Development of the 10-item Edinburgh Postnatal Depression Scale. Br J Psychiatry. 1987 June;150:782–6.

30. Boulund FA, Debelius A, Olsson JC. Lisa . ctmrbio/stag-mwc: StaG v0.7.0. Zenodo; 2023.

31. Köster J, Rahmann S. Snakemake--a scalable bioinformatics workflow engine. Bioinformatics. 2012 Oct 1;28(19):2520–2.

32. Chen S, Zhou Y, Chen Y, Gu J. fastp: an ultra-fast all-in-one FASTQ preprocessor. Bioinformatics. 2018 Sept 1;34(17):i884–90.

33. Wood DE, Lu J, Langmead B. Improved metagenomic analysis with Kraken 2. Genome Biol. 2019 Nov 28;20(1):257.

34. Blanco-Míguez A, Beghini F, Cumbo F, McIver LJ, Thompson KN, Zolfo M, et al. Extending and improving metagenomic taxonomic profiling with uncharacterized species using MetaPhlAn 4. Nat Biotechnol. 2023 Nov;41(11):1633–44.

35. Franzosa EA, McIver LJ, Rahnavard G, Thompson LR, Schirmer M, Weingart G, et al. Species-level functional profiling of metagenomes and metatranscriptomes. Nat Methods. 2018 Nov 30;15(11):962–8.

36. Vieira-Silva S, Falony G, Darzi Y, Lima-Mendez G, Garcia Yunta R, Okuda S, et al. Species-function relationships shape ecological properties of the human gut microbiome. Nat Microbiol. 2016 June 13;1(8):16088.

37. RStudio Team (2020). RStudio Integrated Development for R. RStudio, PBC, Boston. - References - Scientific Research Publishing [Internet]. [cited 2025 Feb 25]. Available from: https://www.scirp.org/reference/referencespapers?referenceid=3542223

38. Wickham H. Easily Install and Load the “Tidyverse” [R package tidyverse version 2.0.0] [Internet]. Comprehensive R Archive Network (CRAN). 2023 [cited 2025 Oct 2]. Available from: https://cran.r-project.org/web/packages/tidyverse/index.html

39. Villanueva R, Chen ZJ. Measurement (Mahwah NJ). Elegant Graphics for Data Analysis. 2019;2:160–7.

40. Allen M, Poggiali D, Whitaker K, Marshall TR, van Langen J, Kievit RA. Raincloud plots: a multi-platform tool for robust data visualization. Wellcome Open Res. 2019;4:63.

41. Haynes W. Benjamini–Hochberg method. In: Encyclopedia of Systems Biology. New York, NY: Springer New York; 2013. p. 78–78.

42. Dixon P. VEGAN, a package of R functions for community ecology. J Veg Sci. 2003 Dec;14(6):927–30.

43. Ruebel ML, Gilley SP, Yeruva L, Tang M, Frank DN, Garcés A, et al. Associations between maternal microbiome, metabolome and incidence of low-birth weight in Guatemalan participants from the Women First Trial. Front Microbiol. 2024 Oct 15;15:1456087.

44. Starke S, Harris DMM, Zimmermann J, Schuchardt S, Oumari M, Frank D, et al. Amino acid auxotrophies in human gut bacteria are linked to higher microbiome diversity and long-term stability. ISME J. 2023 Dec;17(12):2370–80.

45. Culp EJ, Goodman AL. Cross-feeding in the gut microbiome: Ecology and mechanisms. Cell Host Microbe. 2023 Apr 12;31(4):485–99.

46. Shahini A, Shahini A. Role of interleukin-6-mediated inflammation in the pathogenesis of inflammatory bowel disease: focus on the available therapeutic approaches and gut microbiome. J Cell Commun Signal. 2023 Mar;17(1):55–74.

47. Ehlers E, Talton OO, Schust DJ, Schulz LC. Placental structural abnormalities in gestational diabetes and when they develop: A scoping review. Placenta. 2021 Dec;116:58–66.

48. Zhang L, Liu C, Jiang Q, Yin Y. Butyrate in energy metabolism: There is still more to learn. Trends Endocrinol Metab. 2021 Mar;32(3):159–69.

49. Moylan HEC, Nguyen-Ngo C, Lim R, Lappas M. The short-chain fatty acids butyrate and propionate protect against inflammation-induced activation of mediators involved in active labor: implications for preterm birth. Mol Hum Reprod. 2020 June 1;26(6):452–68.

50. Pronovost GN, Yu KB, Coley-O’Rourke EJL, Telang SS, Chen AS, Vuong HE, et al. The maternal microbiome promotes placental development in mice. Sci Adv. 2023 Oct 6;9(40):eadk1887.

51. Qin X, Zhang M, Chen S, Tang Y, Cui J, Ding G. Short-chain fatty acids in fetal development and metabolism. Trends Mol Med. 2025 July;31(7):625–39.

52. Buret AG, Allain T, Motta J-P, Wallace JL. Effects of hydrogen sulfide on the microbiome: From toxicity to therapy. Antioxid Redox Signal. 2022 Feb;36(4–6):211–9.

53. Teigen L, Biruete A, Khoruts A. Impact of diet on hydrogen sulfide production: implications for gut health. Curr Opin Clin Nutr Metab Care. 2023 Jan 1;26(1):55–8.

54. Mello AM, Paroni G, Daragjati J, Pilotto A. Gastrointestinal Microbiota and their contribution to healthy aging. Dig Dis. 2016 Mar 30;34(3):194–201.

55. Cani PD, Bibiloni R, Knauf C, Waget A, Neyrinck AM, Delzenne NM, et al. Changes in gut microbiota control metabolic endotoxemia-induced inflammation in high-fat diet-induced obesity and diabetes in mice. Diabetes. 2008 June;57(6):1470–81.

56. Deehan EC, Mocanu V, Madsen KL. Effects of dietary fibre on metabolic health and obesity. Nat Rev Gastroenterol Hepatol. 2024 May;21(5):301–18.

57. Dreisbach C, Nansel T, Peddada S, Nicholson W, Siega-Riz AM. Dietary sugar and saturated fat consumption associated with the gastrointestinal microbiome during pregnancy. J Nutr. 2024 Nov;154(11):3246–54.

58. Goran MI, Plows JF, Ventura EE. Effects of consuming sugars and alternative sweeteners during pregnancy on maternal and child health: evidence for a secondhand sugar effect. Proc Nutr Soc. 2019 Aug;78(3):262–71.

59. Ramne S, Brunkwall L, Ericson U, Gray N, Kuhnle GGC, Nilsson PM, et al. Gut microbiota composition in relation to intake of added sugar, sugar-sweetened beverages and artificially sweetened beverages in the Malmö Offspring Study. Eur J Nutr. 2021 June;60(4):2087–97.

60. Elms L, Hand B, Skubisz M, Best KP, Grzeskowiak LE, Rogers GB, et al. The effect of iron supplements on the gut microbiome of females of reproductive age: A randomized controlled trial. J Nutr. 2024 May;154(5):1582–7.

61. Beckers KF, Flanagan JP, Sones JL. Correction: Microbiome and pregnancy: focus on microbial dysbiosis coupled with maternal obesity. Int J Obes (Lond). 2025 July;49(7):1421.

62. Poston L, Harthoorn LF, Van Der Beek EM, Contributors to the ILSI Europe Workshop. Obesity in pregnancy: implications for the mother and lifelong health of the child. A consensus statement. Pediatr Res. 2011 Feb;69(2):175–80.

