## Supplemental tables S1-S5 for "Metabolic differences in the maternal gut microbiome precede the birth of large-for-gestational-age infants: A nested case-control study"

### Supplementary information

**Table S1. Participant level variables and how they were scored.**

| **Variable** | **Description** |
|---|---|
| Age | Age in years. |
| BMI ($kg/m^2$) | Body mass index before pregnancy calculated based on self-reported weight and height. |
| Diet score | Score of 0-5 where a higher score indicates a less healthy diet. One point given each for answers other than “daily” to questions about eating fruit, vegetables, and whole grain bread. Additionally, answers of “daily” and “several times weekly” for questions about consumption of sugar and sweetened beverages gives one point each(26). |
| Socioeconomic | A score of 0-3 for socioeconomic status. One point given each for having a bachelor’s degree or higher, being in a relationship and working full time. |
| Smoking | A smoking habit score of 0-2. One point given if ex-smoker or occasional smoking, two points given for regular smoking. |
| Depression score | Measured using Edinburgh postnatal depression scale (EPDS) which contains 10 questions. Each question is scored as 0-3 giving possible scores of 0-30 (26). |
| Stress score | Measured using perceived stress score 4 (PSS-4). Answers to four questions that each award 0-4 points giving possible scores of 0-16. (Cohen S 1983) |
| Antibiotic usage | Does the person take Antibiotic or not. Can be “Yes” or “No”. |
| Alcohol intake (three months before pregnancy) | Does the person take Alcohol or not. Can be “Yes” or “No”. |
| Alcohol intake (during pregnancy) | Does the person take Alcohol or not. Can be “Yes” or “No”. |
| Iron supplements | Does the person take supplements or not. Can be “Yes” or “No”. |

| | |
|---|---|
| Probiotic supplements | Does the person take supplements or not. Can be “Yes” or “No”. |
| Vitamin supplement | Does the person take supplements or not. Can be “Yes” or “No”. |
| Gestational Diabetes Mellitus (GDM) | Whether the participant has a GDM diagnosis or not. Can be “Yes” or “No”. |
| Diet group | If the participant has a specific choice of diet. Can be “plant-based diet” for those who chose “Vegetarian”, “vegan” and “Pescetarian”; Otherwise, considered as “omnivorous”. |
| Food Variables | Fruit, vegetable, whole grain bread, sugar and free sugar drink measured based on more rarely, once a week, several times per week, and daily<br>Daily fiber and fish intake can be “Yes” or “No”. |
| Primipara | Primiparous refers to women experiencing their first pregnancy (26). Primiparous, “Yes” or “No”. |
| Excess weight gain | Excessive gestational weight gain refers to gaining more weight during pregnancy than the amounts recommended by the Institute of Medicine (IOM) based on a woman's pre-pregnancy Body Mass Index (BMI). Excessive weight gain, “Yes” or “No.”(74) |

**Table S2(a): Maternal food frequency intake during gestation, TP1**

| Variables | category | **LGA (245)**<br>N (%) | **AGA (490)**<br>N (%) | P-value* |
|---|---|---|---|---|
| Fruit<br>(frequency consumption) | 1 time/week<br>Daily<br>Several times/week<br>More rarely | 13 (5.31)<br>162 (66.1)<br>63 (25.7)<br>7 (2.86) | 23(4.69)<br>344 (70.2)<br>116(23.7)<br>7(1.43) | 0.4664 |
| Vegetables<br>(frequency consumption) | 1 time/week<br>Daily<br>Several times/week<br>More rarely | 3 (1.22)<br>173 (70.6)<br>67(27.3)<br>2(0.816) | 9 (1.84)<br>365 (74.5)<br>116(23.7)<br>- | 0.133 |
| Fiber rich bread | 1 time/week<br>Daily<br>Several times/week | 58(23.7)<br>39(15.9)<br>92(37.6) | 101(20.6)<br>100(20.4)<br>195(39.8) | 0.276 |

| | | | | |
|---|---|---|---|---|
| (frequency consumption) | More rarely | 56 (22.9) | 93(19.0) | |
| Sugar drinks (frequency consumption) | 1 time/week<br>Daily<br>Several times/week<br>More rarely | 76(31.0)<br>14(5.71)<br>39(15.9)<br>116(47.3) | 150(30.6)<br>22(4.49)<br>80(16.3)<br>238(48.6) | 0.901 |
| Sugar free drinks (frequency consumption) | 1 time/week<br>Daily<br>Several times/week<br>More rarely | 33(13.5)<br>9(3.67)<br>35(14.3)<br>167(68.2) | 91 (18.6)<br>22(4.49)<br>62(12.7)<br>314(64.1) | 0.339 |
| Daily fiber | Yes | 202(82.4) | 425(86.7) | 0.150 |
| Fish consumption | Yes | 226(92.2) | 438(89.4) | 0.269 |
| Probiotic intake | Yes | 38(15.5) | 80(16.3) | 0.859 |
| Iron supplement | Yes | 112(45.7) | 237(48.4) | 0.564 |
| Vitamin supplement | Yes | 226(92.2) | 459(93.7) | 0.376 |

**Table S2(b): Maternal food frequency intake during gestation, TP2.**

| **Variables** | **category** | **LGA (157) N (%)** | **AGA (314) N (%)** | **P-value*** |
|---|---|---|---|---|
| Fruit (frequency consumption) | 1 time/week<br>Daily<br>Several times/week<br>More rarely | 5(3.18)<br>103(65.6)<br>47(29.9)<br>2(1.27) | 11(4.37)<br>164(65.1)<br>63(25)<br>7(2.78) | 0.546 |
| Vegetables (frequency consumption) | 1 time/week<br>Daily<br>Several times/week<br>More rarely | 4(2.55)<br>113(72.0)<br>39(24.8)<br>1(0.637) | 6(2.38)<br>172(68.3)<br>66(26.2)<br>8(3.17) | 0.622 |
| Fiber rich bread (frequency consumption) | 1 time/week<br>Daily<br>Several times/week<br>More rarely | 36(22.9)<br>37(23.6)<br>62(39.5)<br>22(14.0) | 38(15.1)<br>61(24.2)<br>105(41.7)<br>41(16.3) | 0.502 |
| Sugar drinks (frequency consumption) | 1 time/week<br>Daily<br>Several times/week<br>More rarely | 42(26.8)<br>10(6.37)<br>37(23.6)<br>68(43.3) | 80(31.7)<br>9(3.57)<br>52(20.6)<br>104(41.3) | 0.419 |
| Sugar free drinks (frequency consumption) | 1 time/week<br>Daily<br>Several times/week<br>More rarely | 24(15.3)<br>5(3.18)<br>20(12.7)<br>105(66.9) | 33(13.1)<br>11(4.37)<br>54(21.4)<br>147(58.3) | 0.119 |
| Daily fiber | Yes | 128(81.5) | 208(82.5) | 0.309 |
| Fish consumption | Yes | 145(92.4) | 223(88.5) | 0.680 |
| Probiotic intake | Yes | 17(10.8) | 45(10.8) | 0.057 |

| | | | | |
|---|---|---|---|---|
| Iron supplement | Yes | 120(76.4) | 191(75.8) | 0.872 |
| Vitamin supplement | Yes | 130(82.8) | 194(77.0) | 0.399 |

*Based on Chi Square tests

#### Table S3: Alpha Diversity Indices by Time Point and Group

| **Alpha Diversity Indices** | **Time Point** | **95% CI** | **P-value*** |
|---|---|---|---|
| Shannon | TP1 | -0.006, 0.092 | 0.086 |
| | TP2 | -0.068, 0.066 | 0.98 |
| Richness | TP1 | 3.26, 24.90 | 0.01 |
| | TP2 | -7.04, 22.70 | 0.30 |
| Evenness | TP1 | -0.0045, 0.0078 | 0.60 |
| | TP2 | -0.012, 0.005 | 0.44 |

*Based on *t* test

#### Table S4: Within-Group Changes in Microbiome Alpha Diversity from TP1 to TP2 in Groups

| **ΔAlpha Diversity Indices** | **95% CI** | **P-value*** |
|---|---|---|
| ΔShannon | -0.128, -0.008 | 0.025 |
| ΔRichness | -13.389 ,1.909 | 0.1408 |
| ΔEvenness | -0.019, -0.00011 | 0.047 |

Based on *t* test

**Table S5: Group-specific associations between dietary variables and microbiome diversity metric**

| | | **Shannon (LGA vs AGA)** | | | **Richness (LGA vs AGA)** | | | **Evenness (LGA vs AGA)** | | |
|---|---|---|---|---|---|---|---|---|---|---|
| **Predictor** | | Estimate | SE | P* | Estimate | SE | P* | Estimate | SE | P* |
| Sugar drink | TP1 | -0.025 | 0.028 | 0.365 | 0.761 | 6.28 | 0.903 | -0.004 | 0.003 | 0.220 |
| | TP2 | 0.02 | 0.036 | 0.576 | -6.559 | 8.236 | 0.426 | 0.006 | 0.004 | 0.159 |
| Sugar free drink | TP1 | -0.005 | 0.029 | 0.843 | -8.112 | 6.453 | 0.209 | 0.002 | 0.003 | 0.503 |
| | TP2 | 0.04793 | 0.038 | 0.210 | 6.467 | 8.756 | 0.460 | 0.004 | 0.004 | 0.307 |
| Daily fiber | TP1 | -0.033 | 0.069 | 0.633 | -13.324 | 15.384 | 0.386 | -0.001 | 0.008 | 0.872 |
| | TP2 | 0.069 | 0.084 | 0.413 | -11.40 | 19.51 | 0.559 | 0.017 | 0.010 | 0.103 |
| Whole grain bread | TP1 | -0.01398 | 0.025 | 0.579 | 2.811 | 5.572 | 0.614 | -0.003 | 0.003 | 0.241 |
| | TP2 | 0.016 | 0.033 | 0.630 | -0.328 | 7.741 | 0.966 | 0.001 | 0.004 | 0.672 |
| Fruit intake | TP1 | 0.072 | 0.043 | 0.095 | 16.165 | 9.620 | 0.093 | 0.004 | 0.005 | 0.361 |
| | TP2 | 0.016 | 0.058 | 0.773 | -2.256 | 13.125 | 0.8636 | 0.004 | 0.007 | 0.555 |
| Vegetable intake | TP1 | 0.031 | 0.048 | 0.519 | -3.012 | 10.783 | 0.780 | 0.006 | 0.006 | 0.319 |
| | TP2 | -0.043 | 0.06 | 0.473 | -12.256 | 14.071 | 0.384 | -0.002 | 0.007 | 0.773 |
| Fish intake | TP1 | 0.108 | 0.091 | 0.234 | 41.712 | 19.828 | 0.05 | -0.0001 | 0.011 | 0.987 |
| | TP2 | 0.06 | 0.121 | 0.571 | 35.92 | 27.46 | 0.191 | -0.005 | 0.015 | 0.728 |
| Iron (supplement) | TP1 | 0.046 | 0.051 | 0.362 | -0.324 | 11.304 | 0.977 | 0.0081 | 0.006 | 0.202 |
| | TP2 | -0.057 | 0.079 | 0.471 | -18.364 | 18.125 | 0.312 | -0.0009 | 0.01 | 0.925 |
| Probiotic (supplement) | TP1 | 0.009 | 0.07 | 0.889 | 7.424 | 15.506 | 0.632 | -0.0001 | 0.008 | 0.989 |
| | TP2 | -0.0159 | 0.099 | 0.873 | 17.844 | 22.854 | 0.435 | -0.009 | 0.01 | 0.470 |

| | | | | | | | | | | |
|---|---|---|---|---|---|---|---|---|---|---|
| Vitamin (supplement) | TP1 | 0.092 | 0.1006 | 0.357 | 35.87 | 22.17 | 0.106 | 0.0007 | 0.012 | 0.952 |
| | TP2 | -0.06 | 0.085 | 0.451 | -18.323 | 19.615 | 0.351 | -0.003 | 0.0109 | 0.762 |
| Diet group | TP1 | 0.016 | 0.073 | 0.824 | 6.874 | 16.040 | 0.668 | -0.0006 | 0.009 | 0.946 |
| | TP2 | 0.108 | 0.09 | 0.279 | 17.648 | 22.804 | 0.439 | 0.008 | 0.012 | 0.521 |
| Diet score (unhealthy) | TP1 | -0.026 | 0.057 | 0.642 | -8.979 | 12.659 | 0.478 | -0.0001 | 0.007 | 0.980 |
| | TP2 | 0.001 | 0.07 | 0.978 | 3.410 | 16.437 | 0.835 | -0.001 | 0.009 | 0.913 |

*P value based on linear model. P-value<0.05 is significant. SE: Standard Error

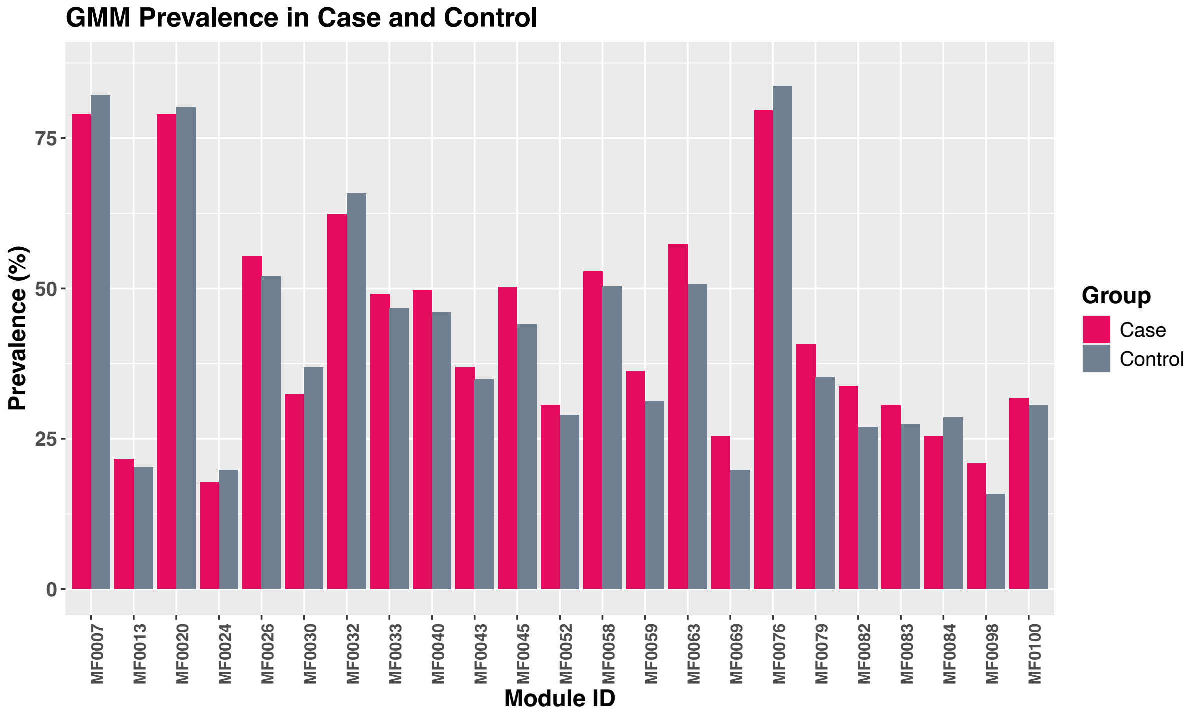


**Figure S1**. Prevalence of intermediate GMM (15–85%) in LGA and AGA groups at TP2 (Chi-square test)
